# A Massively Parallel Screen of 5′UTR Mutations Identifies Variants Impacting Translation and Protein Production in Neurodevelopmental Disorder Genes

**DOI:** 10.1101/2023.11.02.23297961

**Authors:** Stephen P. Plassmeyer, Colin P. Florian, Michael J. Kasper, Rebecca Chase, Shayna Mueller, Yating Liu, Kelli McFarland White, Courtney F. Jungers, Slavica Pavlovic Djuranovic, Sergej Djuranovic, Joseph D. Dougherty

## Abstract

*De novo* mutations cause a variety of neurodevelopmental disorders including autism. Recent whole genome sequencing from individuals with autism has shown that many *de novo* mutations also occur in untranslated regions (UTRs) of genes, but it is difficult to predict from sequence alone which mutations are functional, let alone causal. Therefore, we developed a high throughput assay to screen the transcriptional and translational effects of 997 variants from 5′UTR patient mutations. This assay successfully enriched for elements that alter reporter translation, identifying over 100 potentially functional mutations from probands. Studies in patient-derived cell lines further confirmed that these mutations can alter protein production in individuals with autism, and some variants fall in genes known to cause syndromic forms of autism, suggesting a diagnosis for these individual patients. Since UTR function varies by cell type, we further optimized this high throughput assay to enable assessment of mutations in neurons *in vivo*. First, comparing *in cellulo* to *in vivo* results, we demonstrate neurons have different principles of regulation by 5′UTRs, consistent with a more robust mechanism for reducing the impact of RNA secondary structure. Finally, we discovered patient mutations specifically altering the translational activity of additional known syndromic genes *LRRC4* and *ZNF644* in neurons of the brain. Overall our results highlight a new approach for assessing the impact of 5′UTR mutations across cell types and suggest that some cases of neurodevelopmental disorder may be caused by such variants.

## Introduction

Sequence variation in 5′ untranslated regions (UTRs) can profoundly impact gene expression by perturbing the regulation of translation initiation and efficiency. Variants in 5′UTRs have been implicated in numerous human diseases (Calvo et al., 2009), yet they remain understudied compared to coding variants due to limitations in their capture by exome data and difficulty in interpreting their functional significance. Nonetheless, the contribution of 5′UTR variation to disease has become increasingly well appreciated with the growing availability of whole genome sequences (An et al., 2018; Wright et al., 2021). A recent investigation of variants from 15,000 genome sequences highlighted the severity of single nucleotide variants perturbing upstream open reading frames (uORFs) in 5′UTRs, observing some classes of variants to be constrained to a similar degree as coding missense variants (Whiffin et al., 2020). While limited to variants annotated to impact uORFs, this finding demonstrates certain classes of variation in 5′UTRs can rival the deleterious loss-of-function effects of coding variation.

In addition to uORFs, 5′UTRs contain a variety of *cis*-regulatory elements including RNA binding protein (RBP) binding sites and structured regions which finely tune the translation efficiency of particular transcripts. The functional effects of any of these elements may be sensitive to single nucleotide variants, the start codons of uORFs notably being the most vulnerable class in this regard. The presence of a uORF (predicted by an upstream start codon; uAUG) typically is thought to attenuate the translation of the main open reading frame; however, the functional interpretation of uORF perturbing variants is not often trivial: the consequence of uORF translation on the main ORF depends on local sequence context and positioning relative to the main coding sequence, as in the case of stress-responsive activation of *ATF4* translation (Vattem and Wek, 2004). Furthermore, the effects of variants on higher-order sequence features, such as structured regions, are typically even more opaque to interpretation. Finally, it is unclear to what extent these elements change their functional consequences in distinct cellular contexts and conditions, as different cell types express distinct sets of RBPs.

The challenge of predicting the impact of 5′UTR variants from sequence alone warrants the development of methods that can assay their functional effects at scale, keeping pace with their accelerating rate of discovery. To that end, a number of massively parallel assays have been developed to measure the translation-regulating activity of thousands of 5′UTR sequences. Several methods have quantified the association of short, synthetic UTR sequences with polyribosomes, offering a detailed mechanistic description of key sequence-level regulatory elements, including uORFs and RBP binding sites (Jia et al., 2020; Sample et al., 2019). More recently, efforts have been made to extend the scale of these assays to measure the effect of disease-related variants within the full endogenous sequence context, increasing the likelihood of recapitulating the effect of variants in long, structured UTRs (Lim et al., 2021; Schuster et al., 2023).

High-throughput functional assays are particularly useful for measuring the *in vivo* effects of rare and *de novo* variants, thereby expanding the scope of variant information available to study genetically heterogeneous diagnoses including autism spectrum disorder (autism). Recent whole genome sequencing of autism probands and their families has identified thousands of *de novo* variants(An et al., 2018), complementing work using whole exome sequencing which associated hundreds of novel risk genes with autism by observing an excess of loss-of-function coding mutations in probands(Fu et al., 2022). 5′UTR variants also represent an additional pool of prospective causal variants for autism and related disorders in particular: numerous syndromic neurodevelopmental disorders are caused by loss of function in genes that regulate translation initiation either globally, in the case of *EIF3G*, *PTEN*, and *TSC1/2*, or for a subset of transcripts, as with *DDX3X* and *FMRP* (Calviello et al., 2021; Darnell et al., 2011; Satterstrom et al., 2020; Snijders Blok et al., 2015; Vignoli et al., 2015). Likewise, 5’ UTR mutations in known neurodevelopmental disorder genes *NF2*, *MEF2C* and *SETD5* have now been found in individuals with symptoms consistent with these syndromes (Martin-Geary et al., 2023; Whiffin et al., 2020; Wright et al., 2021). Therefore, to functionally characterize and screen for new variants of interest, we developed a screening strategy to prioritize likely functional and thus potentially causal variants from among 997 *de novo* mutations discovered in autism probands and their families. We first developed and applied a massively parallel assay (MPRA) of translation to measure 5′UTR variants observed in autism probands *in cellulo* (**Fig. 1A,B**). Using this approach we show how measures of translation are impacted by 5′UTR sequences and variation, examining the impact of uORFs and structure. The MPRA results also prioritize a subset of mutations for analysis of protein production, identifying dozens with functional consequences in reporter assays, a subset of which clearly alter allelic abundance in the mRNA, and protein levels in proband cells. Finally, we adapt our MPRA to assess the effects of the mutations in neurons *in vivo,* demonstrating profound differences in sequence-specific regulation across cellular contexts, and identifying additional mutations from individuals with autism that specifically alter translation in excitatory neurons of the maturing brain.

**Figure 1:**
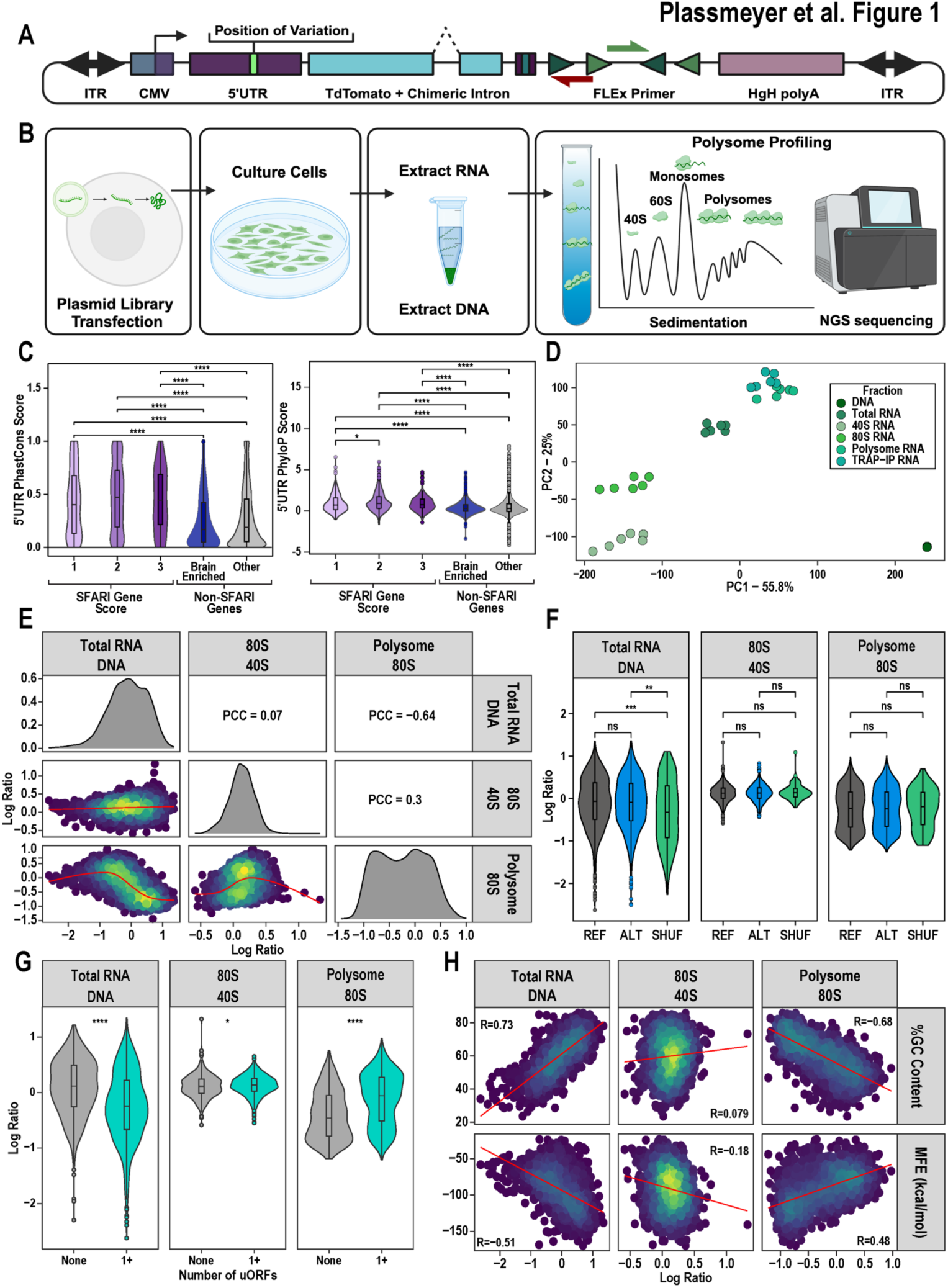
The impact of UTR elements on measures of translation *in cellulo*. **A)** Illustration of the reporter construct showing location of inserted 5′UTR elements, a spliced tdTomato reporter gene, barcode sequence, and RT primer binding site flanked by LoxP sites. **B)** From each biological replicate, total RNA and DNA were collected. A portion of the total RNA was fractionated into 40S-, 80S-, and polysome-associated RNA. **C)** Box plots overlaid on violin plots comparing the distribution of PhastCons (left) and PhyloP (right) scores of the 5′UTR of scored SFARI genes, brain-enriched genes, and other non-SFARI, non-brain enriched genes. Kruskal-Wallis ANOVA and pairwise Wilcoxon tests reveal a significant increase in the conservation scores of SFARI gene 5′UTRs. **D)** PCA plot of MPRA count data showing replicates cluster by fraction type. **E)** Correlograms of the three measures of interest showing significant (*** p < 0.001) positive and negative correlations **F)** Genome-derived 5′UTR sequences (from both variant reference alleles, Ref, and alternative alleles, Alt) display higher transcript abundance compared to a set of randomly shuffled sequences spanning a similar range of GC-content. No difference in 80S/40S enrichment or polysome/80S enrichment between genome-derived and synthetic 5′UTRs. (Mann-Whitney U test: ** p < 0.01, *** p < 0.001) **G)** The presence of uORFs decreases transcript abundance, increases the 80S/40S ratio, and increases polysome/80S enrichment, though with a notably bimodal distribution indicating the impact of a uORF depends on the sequence. (Mann-Whitney U test: * p < 0.05, **** p < 0.0001) **H)** 5′UTR GC content (top row) which corresponds substantially with predicted folding (bottom row, measured by RNAfold-predicted minimum free energy, MFE) positively correlates with RNA abundance and negatively predicts polysome/80S enrichment.

## Results

### A massively parallel assay of translation

To establish the potential for 5′UTR variation to contribute to autism disease risk, we first analyzed the PhastCons and PhyloP conservation scores of the 5′UTRs of genes linked to autism in comparison to the conservation of 5′UTRs in other brain-enriched transcripts (**Fig. 1C**). We discovered that the 5′UTRs of autism-linked genes have increased conservation scores which indicates these genes may be particularly vulnerable to 5′UTR mutations. Therefore, to test whether autism-linked sequence variation in 5′UTRs had a greater likelihood of functional consequences, we sought to functionally characterize the effects of *de novo* mutations identified by whole genome sequencing of 1,902 families from the Simons Simplex Collection (SSC) (An et al., 2018). From 255,106 *de novo* variants discovered genome-wide in SSC probands and matched unaffected sibling controls, we assessed 997 of the 1,163 identified 5′UTR variants with functional screening, determined by which could be synthesized and cloned into our reporter construct. This set of variants included 526 from probands and 471 from sibling controls for annotation and subsequent functional analysis.

The nominal increase in the 5′UTR mutation rate in probands over sibling controls may suggest that a small subset of these mutations are functional and potentially causal. However, in contrast to coding sequence stop-gain mutations (An et al., 2018; Satterstrom et al., 2020) genes harboring 5′UTR variants in probands were no more likely to be constrained (probability of being loss-of-function, pLI > 0.9)(Karczewski et al., 2020) than those from controls (Fisher’s Exact Test, F.E.T., p = 0.4206). Variants from probands and controls were about equally as likely to either introduce or remove uORFs (F.E.T, p = 0.5601), and the prevalence of mutations affecting the context of an uAUG also showed no association to autism (F.E.T., p = 0.2393). Thus, in the absence of measurable case-control differences in the predicted disruption of simple sequence features, we set about interrogating the functional impact of these variants by employing a massively parallel assay of translation.

To measure the functional effects of 5′UTRs on different aspects of translational regulation, we implemented a massively parallel reporter assay (MPRA) which uses the relative association of reporter mRNAs with specific polysome fractions to infer the efficiency of translation initiation and elongation (Cottrell et al., 2018; Lim et al., 2021; Sample et al., 2019). To design pairs of ‘reference’ (the canonical allele) and mutant (the allele found in the family) reporters for each variant, we selected up to 220 nucleotides (nt) of transcript sequence flanking the position of variation, a length that could be synthesized in high throughput and with high fidelity by arrayed oligo library synthesis. This length also corresponds approximately to the median 5′UTR length in the human genome (Leppek et al., 2018). For 286 variants, alternative splicing and transcription initiation sites produced multiple annotated transcript isoforms, creating several non-degenerate sequence contexts. Therefore, all unique transcript contexts were included for each variant. For 579 (38%) reporter sequences, the entire 5′UTR fit within the 220-nt limit for synthesis (**Fig. S1**). The reporter sequence for each allele was tagged with ten unique 10 bp barcodes (**Supplemental Table 1)**. Paired 5′UTR-barcode sequences were cloned after a CMV promoter followed by the insertion of an intron-containing (Younis et al., 2010) tdTomato coding sequence such that the barcode sequence is transcribed in the 3′UTR of the reporter mRNA. To facilitate the later readout of transcript abundance from genetically-defined cell types expressing Cre-recombinase, we included a reverse-transcription (RT) priming site flanked by inverted LoxP sites in the reporter 3′UTR adjacent to the barcode (**Fig. 1A**). Inversion of the Lox cassette by Cre leads to the expression of reporter transcripts which can be selectively reverse-transcribed.

The translational regulatory effects for the final library of 1,507 allelic pairs of 5′UTR reporters were first assayed in HEK cells as a screen to identify alleles that altered translational processes, and thus might also alter protein expression (**Supplemental Table 1**). After harvesting the cells, a portion of the lysate was withheld for estimating steady-state reporter abundance (total RNA), and the remaining lysate was fractionated by density centrifugation. Fractions corresponding to transcripts associated with 40S small ribosomal subunit, monosomes, and polysomes were separately pooled and sequenced (**Fig. S2**). As a potential alternative to polysome fractionation, a portion of each lysate was subjected to translating ribosome affinity purification (TRAP) in parallel (Heiman et al., 2008). Finally, plasmid DNA was sequenced from the extracted DNA to determine the copy number of each barcoded element within the transfected cell population (**Fig. 1B**).

The resulting barcode counts were highly reproducible, with barcode recovery rates exceeding 97% across all libraries and pairwise correlations of barcode counts between replicates exceeding 0.988 (Pearson’s correlation coefficient) (**Fig. S3**). Principal component analysis (PCA) of reporter barcode abundance shows a clear separation between the fractions with the two largest components of variation delineating RNA fractions by their translation status (**Fig. 1D**). Amongst RNA fractions, TRAP RNA barcode abundance most closely correlated with that from the polysome-bound fraction, indicating that TRAP enriches for a distinct sub-population of ribosome-bound mRNA and could serve as an alternative approach to fractionation for translational profiling of a reporter library (**Fig. S4**).

From the sequenced fractions, we defined three primary measures of interest. Normalization of total RNA abundance to relative barcode copy number from the sequenced plasmid DNA provides an estimate of steady-state transcript abundance (total RNA/DNA). Normalizing the abundance in the 80S fraction to abundance in the 40S-bound fraction is interpreted as the efficiency of translation initiation (80S/40S). Finally, polysome enrichment relative to either total RNA or 80S abundance (polysome/total RNA and polysome/80S) was used as a proxy for translational efficiency at steady-state.

Across all reporters, steady-state abundance (total RNA/DNA) was inversely correlated with polysome/80S enrichment (Pearson’s Correlation Coefficient, PCC = -0.64, **Fig. 1E**). This trend runs counter to previous observations of positive correlations between transcript abundance and polysome enrichment (Jia et al., 2020; Lim et al., 2021), which may indicate that the polysome enrichment measurement in this assay does not simply indicate translation efficiency but may also report on transcripts which are stalled during translation and undergoing decay processes. Comparison of polysome enrichment to 80S/40S enrichment shows a marginal positive correlation (PCC = 0.30), indicating that complementary but non-redundant information is obtained by measuring reporter abundance in early and late stages of translation initiation.

To further assess the ability of the assay to capture sequence-dependent variation in transcriptional and translational regulation, we compared the distributions of transcript abundance and translational measures for genome-derived 5′UTR sequences to a set of shuffled UTRs spanning the same range of GC content. While translational measures did not differ between these two sets, reporters with genome-derived 5′UTR sequences had higher transcript abundance than the shuffled counterparts, possibly reflecting the presence of *cis*-regulatory elements which positively regulate transcription (Lim et al., 2021) (**Fig. 1F**).

Notably, the sampling of shuffled 5′UTRs was constrained to excluded uAUGs, while a majority of the genome-derived reporter UTRs (1,315 of 1,507) contain at least one uAUG. Within the set of genome-derived UTRs, the presence of at least one uORF significantly affects both transcriptional and translational measures (**Fig. 1G**). On average, uORFs reduced transcript abundance. A modest increase in 80S/40S enrichment suggests that uORF translation generally impedes efficient initiation, resulting in an accumulation of terminating monosomes on reporter 5′UTRs. Polysome/80s enrichment also increased on average when a uORF was present. This may ambiguously reflect the accumulation of translating ribosomes on the uORF and main ORF or the accumulation of terminating ribosomes in the 5′UTR (**Fig. 1G**). These effects displayed some sensitive to the similarity of the uAUG sequence context to the canonical Kozak sequence(Kozak, 1987a) as well as to the distance at which uORF translation terminated relative to the reporter gene coding sequence. Compared to more proximally terminating uORFs, those terminating further upstream of the main reporter gene start codon resulted in a greater decrease in transcript abundance and greater increase in polysome/80S enrichment (**Fig. S5**).

While impacts of uORFs were complex and sequence context-dependent, sequence features related to RNA structure including GC content and predicted folding free energy robustly predicted transcriptional and translational measurements. The GC content of reporter 5′UTRs had a stabilizing effect on reporter transcript abundance while conversely having a strong negative effect on polysome/80S enrichment and to a lesser extent on 80S/40S enrichment. Correlations with the folding free energy predicted with RNAfold (Hofacker, 2003) follow trends opposite to GC content as expected, albeit with a weaker relationship to each of the measurements (**Fig. 1H**). Overall, GC content alone explains 53% and 46% of the variation in steady-state transcript abundance and polysome/80S enrichment, respectively.

Given the ability of the assay to detect the effect of various features on transcript abundance and translation across a library of natural and synthetic 5′UTRs, we next asked if our method could distinguish the effect of single nucleotide variants (SNVs) and short insertions and deletions (indels). First, to understand the relationship between changes in RNA abundance across different translation state fractions and protein production, we compared the transcriptional and translation MPRA measurements to a dual luciferase reporter assay for a number of controls (**Fig. 2A-D**). To confirm the sensitivity of the MPRA to perturbations in folding free energy, we compared a 5′UTR containing a 15-bp stem-loop to a variant mutated to form no predicted secondary structure (Cottrell et al., 2017). The stem-loop containing 5′UTR drove nearly 20-fold lower reporter expression in the luciferase assay. The MPRA measurements show an increase in transcript abundance yet a two-fold depletion of the stem-loop containing reporter from the polysome fraction relative to the 80S fraction. Thus, for a structural perturbation, the effect detected in the MPRA corresponds to the measured outcome on protein production in the luciferase assay with a notable difference in the magnitudes of the effects, indicating that relatively small changes in polysome fraction enrichment may correspond to larger effects in protein production. Next, we assayed a number of control pairs testing perturbations to uAUGs. An SNV introducing a uAUG initiating an out-of-frame ORF overlapping the reporter gene coding sequence resulted in nearly four-fold lower expression in the luciferase assay. However, the measured effects on transcript abundance and translation in the MPRA revealed more subtle effects, with a subtle shift toward higher polysome enrichment for the uAUG-containing reporter. This result replicated for pairs of controls in which an in-frame, overlapping uAUG shifts to either remaining reading frame (**Fig. S6**). From these controls, it is apparent that small changes in polysome occupancy can correspond to larger changes in protein expression observed in an independent luciferase assay, and similar directional changes in MPRA polysome enrichment can correspond to opposite effects in final protein expression, depending on the 5′UTR. Thus, while the ultimate direction of 5′UTR effects on protein synthesis did not follow simple rules, the MPRA identified perturbations that can change final protein production, and thus could be a useful screen.

**Figure 2:**
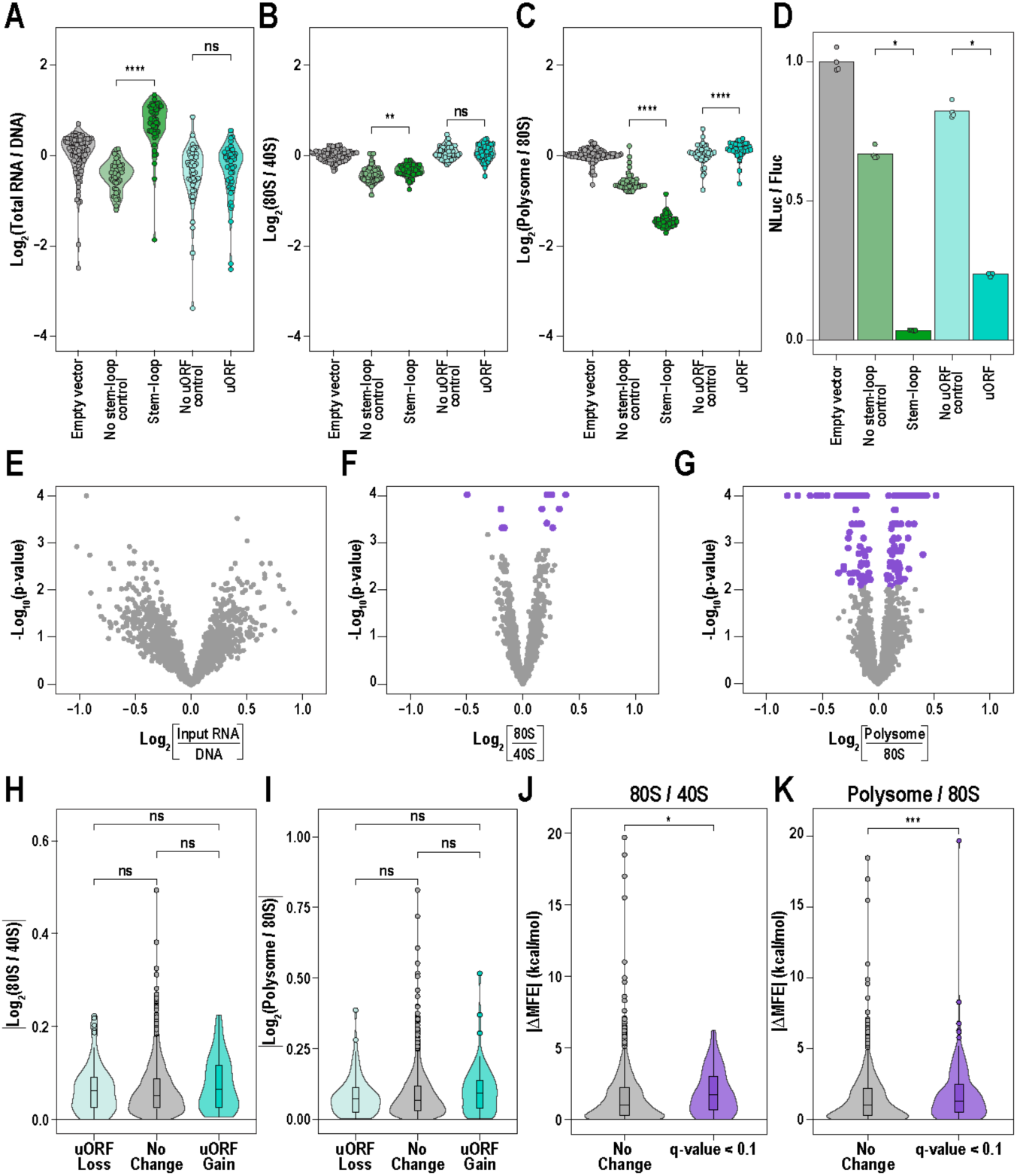
The functional effects of 5′UTR variants primarily impact translation. **A-C)** Synthetic controls demonstrate the sensitivity of different MPRA measurements to changes in secondary structure or uORFs. Compared to a sequence without secondary structure, a synthetic 5′UTR containing a 15-bp stem-loop increases overall transcript abundance and 80S occupancy relative to 40S- or polysome-associated fractions. A 5′UTR containing a short uORF proximal to the reporter start codon has no measurable effect on transcript abundance compared to similar sequence with a disrupted uAUG; however, the uORF slightly increases polysome enrichment relative to the 80S-associated fraction (Mann-Whitney U test: ** p < 0.01, **** p < 0.0001). **D)** A dual luciferase reporter assay shows that both the stem-loop- and uORF-containing 5′UTR sequences result in decreased protein expression despite differences in the response of various MPRA measurements (n=4 per construct, Mann-Whitney U test: * p < 0.05). **E-G)** Testing for allelic effects across 1,507 variant reporter sequences reveals that SNV and short indel variants in 5′UTRs predominantly lead to changes in translation initiation and efficiency rather than transcript abundance (purple points indicate allelic comparisons with q-value < 0.05, see Methods). **H-I**) Across the library, changes in the number of uORFs on average do not result in significantly great allelic effects at the level of translation initiation or translation efficiency (Mann-Whitney U test). **J-K)** Conversely, variants which perturb initiation and translation efficiency (at a threshold of q-value < 0.1) are predicted to have greater changes in folding free energy (Mann-Whitney U test: * p < 0.05, *** p < 0.001).

### Functional 5′UTR variants primarily impact translation initiation

Next, we assessed the functional impacts of variants from probands and controls on transcript abundance and translation in HEK cells. 5′UTR variants were found far more likely to perturb translation regulation rather than transcript abundance. Of the 997 variants, none were found to significantly alter transcript abundance (q-value < 0.05). Conversely, 154 altered polysome/80S or polysome/total RNA enrichment in at least one transcript isoform context. Only 13 variants altered 80S/40S enrichment, and of these, 6 also perturbed polysome/80S or polysome/total RNA enrichment (F.E.T., p = 0.008) (**Fig. 2E-G, Supplemental Table 2**). In general, the log2 fold changes measured across transcriptional and translational effects were only weakly correlated, with effects on transcript abundance sharing a slight negative correlation with effects on polysome/80S enrichment (PCC = -0.23, p = 6x10^-19^), suggesting that variants tend to have largely orthogonal effects on transcription abundance and regulation of translation (**Fig. S7**).

We next evaluated the effects of the variants when considering their predicted effects on uORFs, sequence motifs, and structure. First, we asked whether mutations resulting in the gain or loss of a uORF through the disruption of either a uAUG or upstream stop codon tended to have a greater effect on measures of translation. Neither the grain nor loss of a uORF was associated with larger effect sizes on average relative to other variants for any measure of translation (**Fig. 2H-I**). Next, we asked if variants that perturb sequence motifs tended to have larger effect sizes. The gain or loss DNA binding motifs (Weirauch et al., 2014) resulted in larger shifts in transcript abundance on average (Mann-Whitney U test, p = 0.002); however, variants that perturbed RBP binding sites (Ray et al., 2013) did not result in greater shifts in any translational measure (**Fig. S8**). Consistent with the strong correlation of transcriptional and translational measures with predicted RNA folding energy observed across elements, we found that functional variants affecting either 80S/40S enrichment or polysome/80S enrichment on average had larger predicted changes in the folding free energy of the reporter 5′UTR (**Fig. 2J-K**).

Finally, we tested if *functional* 5′UTR mutations might be a common contributor to autism, in which case a greater fraction of such mutations would be present in the probands compared to their siblings. However, an enrichment of functional proband variants was not observed. Only 77 of the 161 variants perturbing 80S/40S, polysome/80S, or polysome/total RNA enrichment were observed in probands which account for 526 of the 997 assayed variants (F.E.T. for proband enrichment, p = 0.89). While we did not have an *a priori* hypothesis about the direction of effect, functional proband variants were not associated with a consistent direction of effect on any of the translational measures, nor were they more likely to perturb ‘constrained’ genes (F.E.T. for enrichment of functional proband variants in genes with pLI > 0.9, p = 0.86), which are frequently mutated with stop-gains in autism. While an excess of loss-of-function protein-coding variants has been previously reported amongst autism probands (Satterstrom 2020), 5′UTR variants may only carry a less severe burden for autism and thus will likely be a rarer cause of autism than stop-gain mutations. Yet, overall, our assay uncovered 77 proband variants in total with functional effects on translational regulation (**Supplemental Table 2**).

### A subset of UTR variants also impact protein production

In total, 16% of the variants showed a measurable effect on the relative association with the 40S, 80S, or polysome fractions. We next tested a subset of these to determine the rate at which the MPRA identified variants that also had the potential to alter protein expression using a dual luciferase reporter assay in HEK cells. We selected variants from probands that affect genes for which there is evidence of association with autism (SNX5, *NR4A2*, *MYO9B*, *GABRB3*, *LRFN5*, *ZMYND11, C12orf57, PCCA*) (SFARI score ≤3, or S)(Abrahams et al., 2013). An additional 13 proband variants were selected which affected constrained genes (pLI > 0.9) that were found to be expressed in GTEx (version 8) (GTEx Consortium, 2020) brain tissues above a threshold of 0.1 transcripts per million (*USP48*, *ELOA*, *HSPA8*, *PPP1CC*, *PPP1R12A*, *EFNB2*, *CLIP3*, *GMEB2*, *OXSR1*, *PBRM1*, *CALD1*, *MKRN1*, *SMC5*). Lastly, we included the variants with largest effects for any of the transcriptional or translational measures from the MPRA, drawing from both the proband and sibling control mutations (see **Supplemental Table 3** for full details on variants tested by luciferase assay). In total, 34 functional variants with a translational effect in the MPRA were selected along with an additional 17 variants with similar characteristics which had no effect in the MPRA.

Of the 51 variants assayed, 31 significantly altered protein expression (12 increased and 19 decreased, FDR-corrected Mann-Whitney p < 0.05) (**Fig. 3A**). While a majority of these changes had small effect sizes, 7 variants altered expression by over 50% with the largest magnitude effects arising from variants that increased expression. Several MPRA measures predicted which variants altered protein expression, though not with perfect specificity. Of the four sampled variants that altered 80S/40S enrichment, only one altered protein expression. Yet for the 29 assayed variants that altered polysome/80S enrichment, 18 had an effect at the protein level. In general, the luciferase expression and the difference between alleles did not correlate with the 80S/40S enrichment, polysome/80S enrichment, or transcript abundance measurements (**Fig. S9**), consistent with our results establishing the assay (**Fig 2A-D**). Thus, though many MPRA-identified variants also alter protein levels, the relationship between MPRA results and magnitude or direction of protein effect does not follow simple rules.

**Figure 3:**
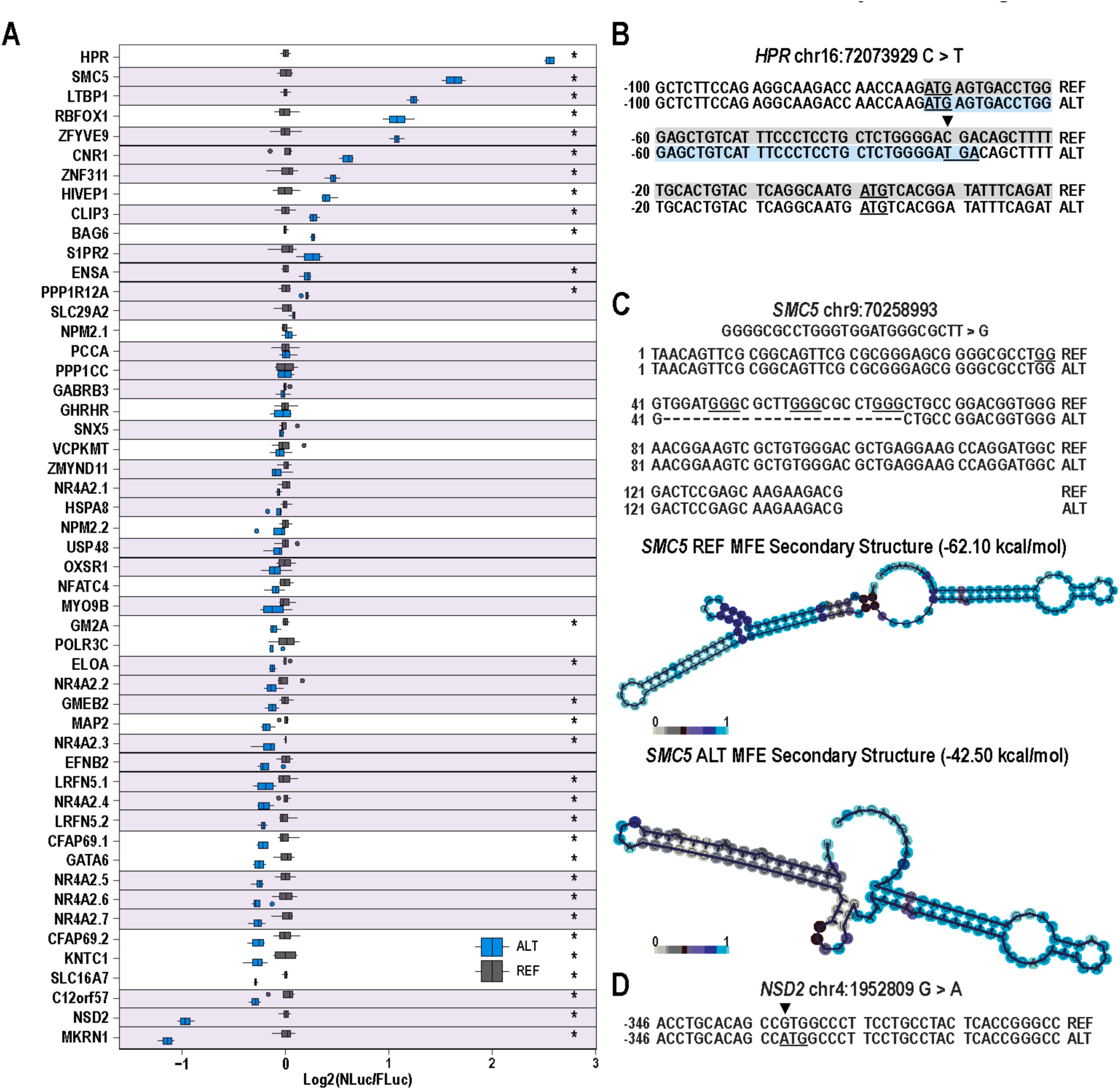
5′UTR variants identified by MPRA also alter protein production in reporter assay. **A)** A dual-luciferase reporter assay performed on a sampling of 51 variant reporters reveal numerous effects on protein expression (n= 4 per experiment, * = FDR-adjusted p < 0.05, Mann-Whitney U test). For presentation purposes, the log-normalized luciferase signal is adjusted to the reference allele within each pair. Purple highlight/outline represents cases while unhighlighted rows show controls. **B)** A C>T transition variant from an unaffected sibling in the 5′UTR of *HPR* introduces a stop codon downstream of an uAUG, allowing for termination of translation of a uORF and subsequent reinitiation to rescue translation of the main ORF, leading to a nearly 6-fold increase in reporter expression. **C)** A deletion variant from a proband in the 5′UTR of *SMC5* removes a series of guanine repeats (underlined) predicted to form a G-quadruplex <u>(Kikin et al., 2006)</u>. The truncated UTR is predicted to have a reduced minimum free energy of folding as predicted by RNAfold <u>(Gruber et al., 2008)</u>. **D)** A G>A transition variant from a proband in the 5′UTR of *NSD2* introduces an uAUG, leading to decreased expression of the luciferase reporter encoded by a downstream out-of-frame ORF.

An examination of the variants with the largest effect revealed diverse mechanisms underlying translational perturbation. A sibling variant observed in the 5′UTR of *HPR* had the largest effect size, resulting in a 6-fold increase in reporter expression (**Fig. 3A,B**). A stop codon introduced by this SNV is predicted to terminate translation of an overlapping upstream open reading frame (oORF) that may otherwise inhibit expression of the reporter gene. The next largest effect came from a proband variant in the 5′UTR of *SMC5*: a 25 bp deletion at chr9:70258993 which led to a 3-fold increase in luciferase expression. This effect likely results from derepression of translation initiation through relaxation of secondary structure in the 5′UTR. The nucleotides deleted in this mutation are predicted to form a G-quadruplex structure or otherwise contribute to an extended hairpin structure (Gruber et al., 2008; Kikin et al., 2006) (**Fig. 3C**). A second large effect proband variant, chr4:1952809G>A located in the 5′UTR of *NSD2*, resulted in a 50% decrease in luciferase expression through the introduction of an uAUG that initiates an oORF (**Fig. 3D**). Thus, several variants of large effect in reporter assays were detected from proband genomes.

### UTR variants can also alter translational features of the host gene and protein production by proband cells

Limitations on the length of constructs used to screen variants in both of the reporter assays potentially exclude important sequence context and positional information that may attenuate variant effects within the native transcript. Therefore, we took advantage of existing patient lymphoblastoid cell lines (LCLs) available from the SSC to examine differences in expression and translation of transcripts from the variant alleles. We hypothesized that if the variant indeed alters translation in the native genomic context, we should see altered counts of RNA molecules containing the variant allele relative to the WT allele following polysome fractionation — i.e. an “allelic translation imbalance” (ATI) study, analogous to allelic expression imbalance (AEI) (Chuang et al., 2017; Liu et al., 2020; Mascarenhas et al., 2015; Mohammadi et al., 2019). To control for PCR bias when amplifying loci of indels, we further normalized allelic counts from total RNA samples against counts from genomic DNA sequencing which would share any such technical bias. From the set of variants with functional effects in both the MPRA and luciferase assay, we selected six for which expression of the relevant host gene isoform in LCLs was confirmed as expressed by RT-PCR.

Of the six variants assessed for AEI and ATI, two (occuring in *MKRN1* and *USP48*) showed concordance with the corresponding MPRA measurement in direction and magnitude (**Fig. 4A-D**). Across both experiments, we found that the indel chr7:140478819 G>GGGGA in the 5′UTR of *MKRN1* reduced relative polysome/80S enrichment by 14% in the HEK cell MPRA while the variant in the full transcript context led to a 16% decrease. The SNV chr1:21783300 A>T in the 5′UTR of *USP48* similarly showed concordance between measurements of polysome/80S enrichment, with a 9% and 31% decrease measured from the MPRA and LCL sequencing, respectively.

**Figure 4:**
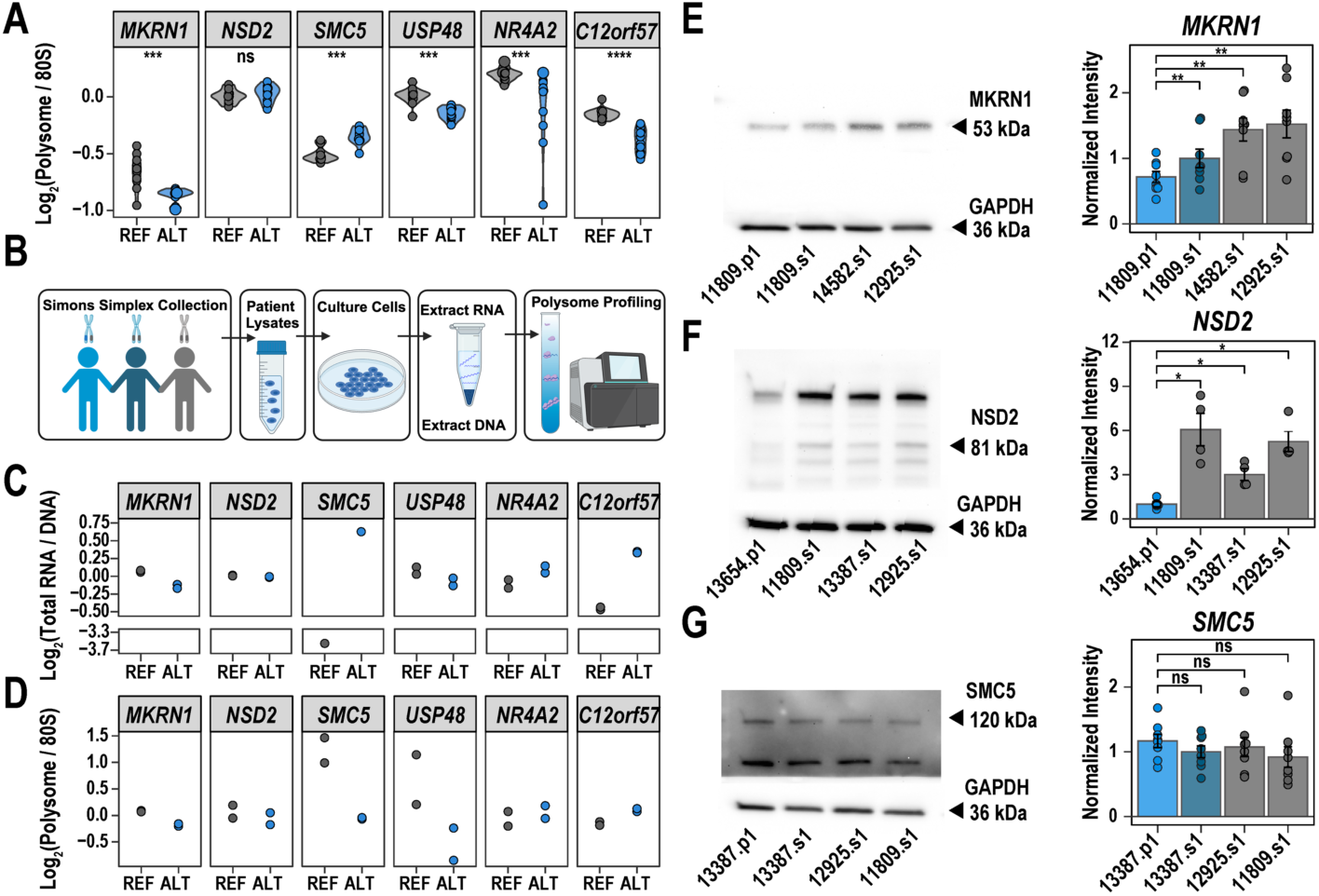
5′UTR variants from autism probands alter translation in the native genomic context. **A)** Examples of polysome/80S enrichments altered by the alternate allele from the MPRA. **B)** LCLs from these patients were harvested and subjected to polysome fractionation followed by targeted sequencing of the variant loci to quantify relative reference and alternative allele abundance. **C)** Differences in the relative abundance of each allele between total RNA and DNA (allelic expression imbalance AEI) was tested, finding that variants in MKRN1, SMC5, and C12orf57 had an effect on transcript abundance. **D)** Similarly, differences in allele abundance between the polysome fraction relative to 80S abundance (allelic translation imbalance, ATI) showed that variants in SMC5, MKRN1, and USP48 had translational effects. (n=2, significance of change in alternative allele abundance between fractions assessed by negative binomial generalized linear model fit, see Methods and Supplemental Table 4). **E-G)** Western blotting of MKRN1, NSD2, and SMC5 in proband-derived cell lines (light blue) alongside cell lines from matched sibling control (dark blue) and cell lines from unrelated control individuals in the Simons Simplex Collection (grey) (n = 8 technical replicates per sample in **E** and **G**, n = 4 technical replicates per sample in **F**; Wilcoxon signed-rank test in **E** and **G**; Mann-Whitney U test in **F**; ** p < 0.01).

Several variants displayed translation effects on full-length transcripts in LCLs that differed from what was measured in the MPRA. A SNV in *NSD2* (chr4:1952809 G>A) resulted in an increase in polysome/total RNA enrichment in the MPRA which was not observed for the transcript in LCLs; however, transcripts from the variant allele showed greater 80S/40S enrichment in the patient cell line (**Fig. S10**). Similarly, an SNV in the 5′UTR of *C12orf57* (chr12:6944014 T>G) altered polysome/80S and polysome/total RNA enrichment in the MPRA which did not replicate in LCLs, yet 80S/40S enrichment was reduced for the variant transcript (**Fig 4D, Fig. S10**). The indel in *SMC5* increased polysome/80S and polysome/total RNA enrichment in the MPRA and drove increased luciferase expression. In the native transcript, this variant drove large effects in the opposite direction of the MPRA (**Fig. 4D, Fig. S10**). Finally, a proband variant in *NR4A2* (chr2:156330692 C>T), which drove decreased polysome/80S and polysome/total RNA enrichment in the MPRA in multiple isoform sequence contexts, had no effect on ATI or AEI in LCLs (**Supplemental Table 4**).

For all six variants screened for ATI and AEI in LCLs, we sought to determine if they altered protein levels in the patient cells compared to individuals not carrying the mutation. As many of these genes are poorly characterized, only three had usable antibodies. For these three, we observed that the patient mutations in *MKRN1* and *NSD2* alter protein production (**Fig. 4E-F, Fig. S11**). In spite of robust changes observed in the luciferase assay and ATI, overall protein level of SMC5 appeared normal (**Fig. 4G, Fig. S11**), suggesting some post-translational buffering of this protein may occur to compensate for potential AEI/ATI effects, at least at steady state in LCLs.

### A cell-type specific MPRA measures altered translation activity *in vivo*

Our results in cell lines indicated that a subset of 5′UTR mutations from patients can alter protein production of host genes. However, regulation of translation can be cell type-specific (Cottrell et al., 2018; Lagunas et al., 2023), as certain factors such as the expression of helicases, initiation factors, and other RNA-binding proteins can vary between cellular contexts. Developing neurons are likely the most important cell type for autism (Fu et al., 2022; Li et al., 2023; Satterstrom et al., 2020). Therefore, we have previously adapted an MPRA for cell type-specific measures in neurons *in vivo* (Lagunas et al., 2023). Our previous approach was not sensitive enough to detect the effect of variants due to technical limitations of measuring small effect sizes with relatively few barcodes per reporter. Here, we increased the number of barcodes and introduced a novel reporter construct with a Cre-inducible switch for recording the cell type of origin of reporter transcripts. With these modifications, we tested whether it was feasible to improve the MPRA in glutamatergic cortical neurons *in vivo* and to detect the effect of individual variants. We targeted developing excitatory cortical neurons which show enriched expression of known autism genes and a time point (postnatal day 21, P21) corresponding to a critical period in circuit maturation for cortical development during the second peak of autism-associated gene expression (Willsey et al., 2013). Such experiments would allow us to test the hypothesis that distinct principles guide translation regulation of reference 5′UTRs *in vivo* compared to what was observed *in cellulo* and to screen for additional patient variants that might be functional specifically in neurons, and thus have potential to be causal for autism.

To avoid a bottleneck in efficient viral transduction of our library, we divided our variant reporters into three sublibraries of approximately 500 allelic reporter pairs. Each sublibrary was mixed with a separately packaged pool of control reporters and delivered transcranially into the cortex of P1 neonates from a cross between Vglut2-Cre^+/-^ and Ef1a:LSL-eGFP-RPl10a^+/-^ (**Fig. 5A**). Six replicate animals genotyped as positive for both the Cre and eGFP-Rpl10a transgenes were used for each sublibrary. Driven by a strong constitutive CMV promoter, the reporter mRNA is expressed by most brain cell types. However, in Vglut2-positive glutamatergic neurons, the expression of Cre inverts the reverse transcription (RT) primer cassette in the reporter 3′UTRs and also removes an early stop codon from the constitutively expressed eGFP-Rpl10a transgene in the genome, labeling the target neuron population for microscopy (**Fig. 5B,C**). Using RT primers complementary to the flipped orientation of this cassette, mRNA abundance can be measured specifically from Vglut2-positive neurons as the cassette has flipped to the “CreON” state, separately from all other transduced cells in which the cassette remains in the “CreOFF” state (**Fig. 5B, Fig. S12**). Total RNA and DNA along with 80S- and polysome-associated mRNA collected from whole brain homogenate was prepared into sequencing libraries using both the CreON and CreOFF state primers (**Fig. S13**). We observed generally robust correlations between replicates of each fraction, with most pairwise Pearson’s correlation coefficients exceeding 0.90 (**Fig. S14**). Notably, barcode recovery (above a threshold of 20 UMI counts set for hypothesis testing) and replicate correlations were particularly low for 80S libraries prepared with the CreOFF RT primer, leading us to exclude measurements using CreOFF state 80S counts from subsequent analyses. As in the HEK MPRA, PCA of barcode counts showed RNA samples clustered by fraction type. Barcode counts also correlated well across CreON and CreOFF libraries with RNA libraries showing marginally lower correlation than DNA abundance (**Fig. S15**).

**Figure 5:**
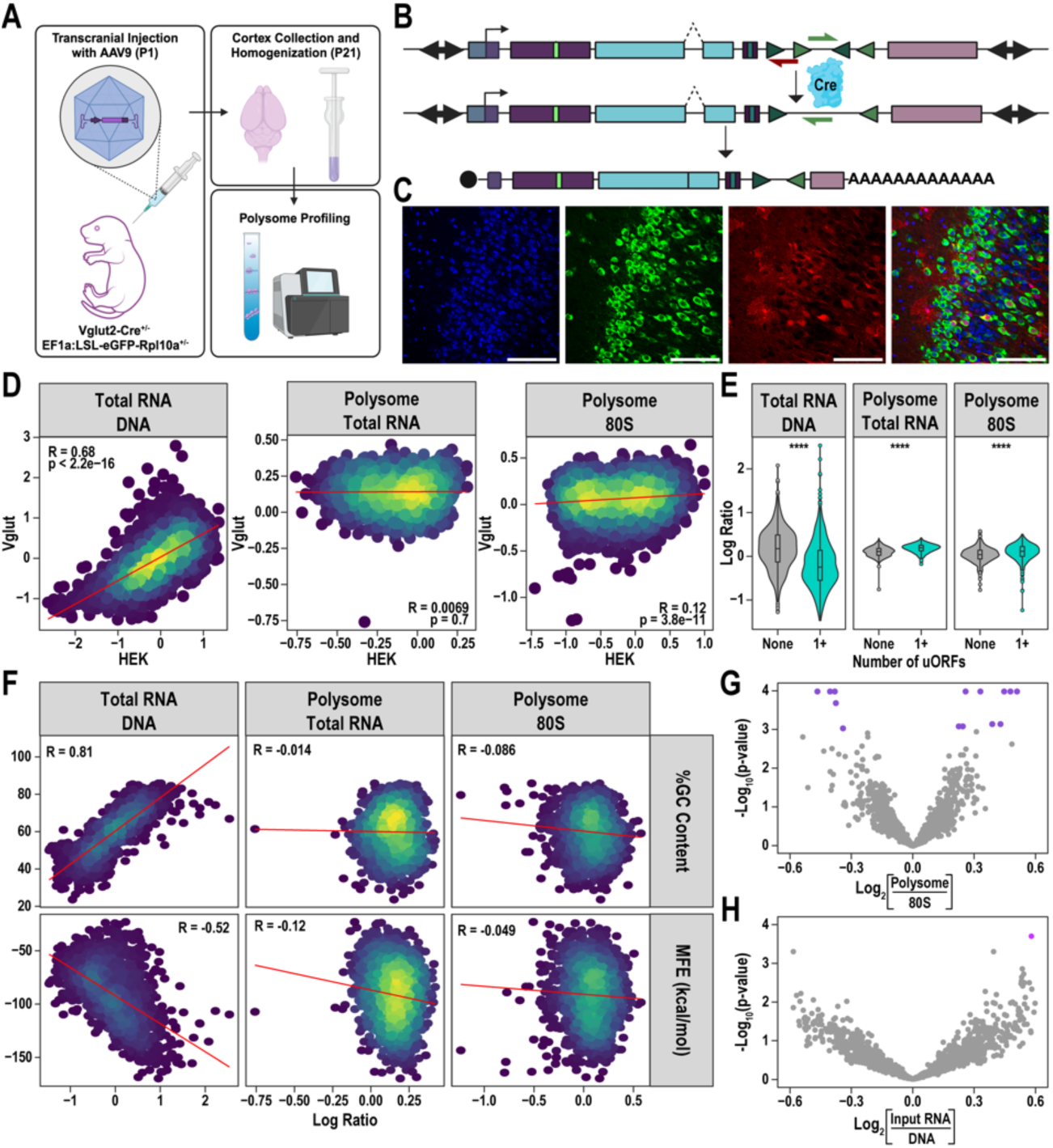
The impact of UTR elements on measures of translation in excitatory neurons *in vivo*. **A)** Illustration of experimental design: the same 5’ UTR libraries were packaged into AAV and delivered into the perinatal mouse brain, collected at P21, and fractionated as above. **B)** LoxP sites enable a Cre-dependent inversion of a primer site (top rows), resulting in mRNAs with a unique RT-primer site in excitatory neurons. **C)** Neuronal and non-neuronal cell types in the lateral cortex display expression of the AAV-delivered tdTomato reporter gene while eGFP expression in Vglut2-positive Cre expressing neurons partially overlap the transduced cell population. (Scale bar: 100 µm) **D)** Comparison of transcript abundance and translation measures between assays performed in HEK and cortical neurons indicate that translational regulation varies substantially between cellular contexts despite relatively similar transcript levels. **E)** For transcripts translated in glutamatergic neurons, the presence of uORFs generally leads to reduced transcript abundance but slightly increases the abundance of transcripts in polysome-bound fractions (Mann-Whitney U test: **** p < 0.0001). **F)** GC content and predicted folding free energy of transcript 5′UTRs strongly predicted transcript abundance but exhibited little correlation with translation measures, unlike translational activity in HEK cells which were well correlated with these features. **G-H)** Testing for allelic effects on reporter transcripts expressed in glutamatergic neurons reveals 6 variants with an effect on transcript abundance and 14 with an effect on polysome-80S enrichment (purple points indicate allelic comparisons with q-value < 0.05).

First, to examine the general principles of translation regulation by 5′UTRs in excitatory neurons *in vivo, w*e compared the log-abundance ratios for reference elements between the *in cellulo* and *in vivo* assays. This supported the hypothesis that translation activity depends highly on the cellular context and that neurons have distinct rules: Despite correlated reporter transcript abundance across both experiments (PCC = 0.68), measures of polysome enrichment relative to total mRNA or 80S-bound mRNA show effectively no correlation between experiments (**Fig. 5D**). Similarly, across the two contexts assayed *in vivo*, reporter transcript abundance was relatively similar between CreON and CreOFF, yet translational measures were less well-correlated (though still showing greater agreement than with the HEK experiment) (**Fig. S16**). Additionally, transcript abundance and translation measures spanned a narrower dynamic range in the *in vivo* assay, and reporter abundances shared higher correlations across each DNA and RNA fraction as well. This decrease in differential partitioning across each fraction *in vivo* may suggest that the activity differences *in cellulo* are buffered *in vivo* (**Fig. S15**).

While some of the effects of key sequence features that drive variation in translation *in cellulo* replicate *in vivo*, there are notable exceptions. The presence of uORFs generally has the same effect on both transcript abundance and translation across both assays, with a pronounced decrease in abundance and subtle increase in polysome-enrichment for reporter UTRs containing a uORF (**Fig. 1F**, **Fig. 5E**). For elements containing a single uORF, we assessed how the strength of the uAUG context (as defined in Whiffen et al. 2021, see Methods) influenced the measured effect on translation and transcription, finding that transcriptional and translational measures were sensitive to the uAUG context of uORFs in both assays (**Fig S5, Fig. S17**). Re-initiation of translation of coding sequences following uORFs depends on the scanning distance and time between the uORF stop codon and coding sequence start codon (Kozak, 2001). Therefore we examined an effect of this distance and found that transcript abundance decreased for uORFs terminating further from the reporter coding sequence in both assays. However, while more distal uORFs correlated with greater polysome-enrichment in the HEK experiment, no such effect of uORF position on translation was observed *in vivo* (**Fig S5, Fig. S17**). The efficiency of re-initiation can depend on the abundance of initiation factors — which can vary by cell type and cell state — and can be impeded by long or structured sequences between the uORF and coding sequence (Bohlen et al., 2020; Calvo et al., 2009; Kozak, 2001). The greater positional dependence of uORFs observed in HEK may therefore suggest differences in re-initiation efficiency compared to the *in vivo* experiment. Finally, we examined the effect or uORF length — which can also affect the rate of downstream re-initiation (Kozak, 1987b) — and found only an effect for decreasing polysome/80S enrichment in glutamatergic neurons *in vivo* (**Fig S5, Fig. S17**).

In addition to differences in uORF regulation, the effects of RNA folding stability and GC content notably differed between experiments. In both contexts, transcript abundance is correlated with GC-content and is anti-correlated with folding free energy; however, the correlations between polysome enrichment and either GC content and folding free energy observed *in cellulo* (**Fig. 1G**) are absent *in vivo* (**Fig. 5F**). While they accounted for a majority of the variation in polysome enrichment *in cellulo*, the absence of this effect *in vivo* results in monosome and polysome loading driven primarily by the underlying transcript abundance of each reporter. Overall, our examination of the principles by which sequence alters 5′UTR function suggests that neurons may have more efficient mechanisms for resolving 5′UTR structure and uORF position than HEK cells.

Finally, we examined the effect of individual mutations on translation in the MPRA. Of the variants measured with sufficient depth in glutamatergic neurons (CreON), two had a significant effect on transcript abundance, six had an effect on polysome/total RNA enrichment, and 15 had an effect on polysome/80S enrichment (q-value < 0.05). From the CreOFF measurements of total RNA and polysome abundance, nine variants had an effect on transcript abundance, only one of which also had an effect in the glutamatergic neurons, while three entirely unique variants were found to affect CreOFF polysome/total RNA enrichment (**Fig. S18**). Testing for an interaction of allele effects with cellular context found 22 and 17 reference alleles that differed in transcript abundance and polysome/total RNA enrichment, respectively, between CreON and CreOFF but found relatively few cases of statistically significant differences in the effect of the variant across contexts (**Fig. S18**, see **Supplemental Table 5** for full *in vivo* results).

Across all 23 variants with significant effects in glutamatergic neurons, 9 were from probands, again indicating no enrichment of probands amongst functional variants (F.E.T., p = 1). Most of the functional proband variants fall in genes that are unlikely to be functionally constrained (pLI < 0.9) or are not expressed in adult brain tissues. However, two variants were identified in autism associated genes: one in *LRFN5* (SFARI score 2) leading to a decrease in polysome/total RNA enrichment and another in *LRRC4C* (SFARI score 1) leading to an increase in polysome/monosome enrichment. A third variant resulting in decreased polysome/total RNA enrichment was identified in *ZNF644* which has strong association with developmental delay (TADA-DD FDR <0.005)(Fu et al., 2022), though not autism specifically. Most of the functional proband variants identified in glutamatergic neurons were not discovered in our initial screen in HEK cells, and in general, variant effects were not correlated across the two experiments (**Fig. S19**). An exception to this was the *LRNF5* variant which appeared as a hit in both assays. For this variant, we utilized two isoform specific sequence contexts and differing reporters. In HEK cells we see decreased polysome/80S enrichment in one sequence context, and for the other context, we see decreased polysome/total RNA enrichment in glutamatergic neurons. In our luciferase assay, the variant results in a small effect size decrease in protein expression in both sequence contexts (**Fig 3A**). Together, these results demonstrate that functional effects on transcript abundance and translation regulation can be screened for hundreds of variants *in vivo*. Deviations in these effects between *in vivo* and *in cellulo* models highlight the significant role cellular context plays in detecting functional effects on translation, underscoring the value of assays compatible with measurement in live animals.

## Discussion

In this work we developed a 5′UTR assay to characterize how different cell types respond translationally to 5′UTR sequences and to systematically screen for allelic effects of 997 *de novo* mutations from autism probands and their siblings. With regard to insights into general principles for how 5′UTRs might influence translation, we had several observations based on analysis of both element effects, and the consequences of the allelic mutations. *In cellulo*, we saw that 5′UTR elements could influence all aspects of post-transcriptional regulation from RNA levels to 40S, 80S, or polysome occupancy, but the relationship between these measures differed *in vivo* in glutamatergic neurons. For example, elements had a similar impact on transcript abundance in both cell types, but while transcript abundance was generally negatively correlated to polysome/monosome enrichment in HEK cells, it was uncorrelated in the neurons. If polysome/monosome enrichment is a proxy for translation, this suggests in HEK cells translation may lead to greater transcript degradation, while in neurons it would not. This could be consistent with differences in NMD pathways in HEKs and other immortalized lines (Gerbracht et al., 2017). Of course, an alternative explanation is that in neurons the polysome/monosome measure may reflect transcripts stalled for transport (Darnell et al., 2011), during which they might be relatively protected. Similarly, GC content/5′UTR structure had less influence on translation neurons and likewise variants altering uORFs and structure in HEK cells had larger effects, while neurons appeared to have more mechanisms for reducing these effects. Thus, across numerous observations, our experiments highlight the importance of cellular context in influencing the role specific 5′UTR elements have on measures of translation.

In seeking to identify which allelic variants might be functional, and thus have potential to be causal, we developed an experimental series to prioritize mutations by function, organized by throughput (**Fig. 6**). The *in cellulo* MPRA assay revealed 161 functional allelic effects from both probands and siblings in roughly equal proportions. In contrast to stop-gain mutations in brain expressed genes (which are roughly twice as frequent in probands(Sanders et al., 2015)), there was only a very slight and non-significant enrichment of functional probands variants in brain expressed genes (∼10% more). This indicates that at the high end, we might expect 5-10 out of 997 (0.5-1%) of the 5′UTR variants to be causal for autism. To further filter and annotate these results, we selected 34 functional hits from the MPRA for assessment by luciferase assay to test for impact on protein production, of which >50% (20/34) had a clear result. Notably, while this was a substantial fraction, the direction of effect was difficult to predict from the MPRA alone, suggesting polysome occupancy does not simply reflect translation of the primary ORF, but could be measuring a mix of uORF translation and stalling as well.

**Figure 6:**
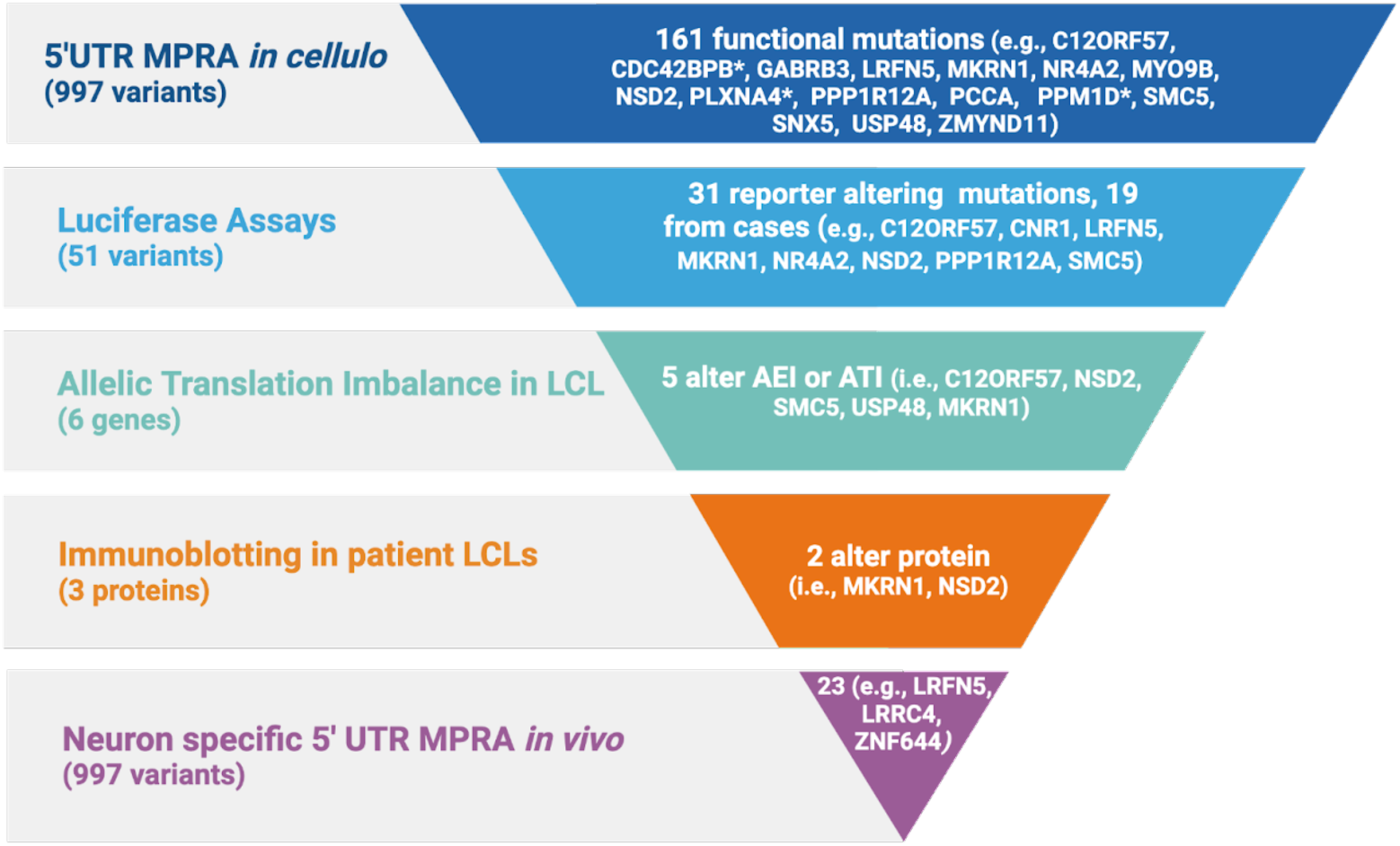
Summary of 5’UTR functional findings. Our *in cellulo* MPRA screened 997 variants from autism probands and sibling controls, finding 161 with a functional consequence on transcript abundance or translation (at q-value < 0.05; 238 variants with q-value < 0.10). A selection of proband variants from constrained genes and known neurodevelopmental disorder genes are highlighted at each stage (with those significant at q-value < 0.10 indicated by *). Luciferase assays from 51 variants selected from both probands and siblings identify 31 that significantly alter protein expression. Of these, 19 variants were from probands and include several in genes with some evidence for association with developmental disorder. Five of these also alter translation from the endogenous locus when measured in LCLs, and two showed significant difference in levels of endogenous protein expression. An additional neuron-specific MPRA identified another 24 mutations showing function in neurons of the developing brain, including three in known neurodevelopmental disorder genes.

Nonetheless, we selected a subset of these for further analysis (based on effect size and predicted expression in LCLs) to determine if the allele also altered features of translation in patient cells using an AEI/ATI strategy in the native context, using the reference allele as an endogenous control, and amplification from genomic DNA as a control for PCR biases. The advantage of this AEI/ATI is that it assesses the effect of the allele in the native sequence context and at endogenous expression levels, but it can only be applied to transcripts and their isoforms expressed in LCLs of the six proband lines selected for analysis. Of these, five variants showed some imbalance in either ATI or AEI, with two (in *MKRN1* and *USP48*) showing concordant effects with the MPRA.

We also attempted to screen four of these probands for effects on the native protein via immunoblot. This is a challenging experiment, given the relatively uncharacterized nature of these proteins and their antibodies, the expectation of potentially small overall effects (since the immunoblot will detect the sum of both the wild type and mutant alleles, but only the mutant allele is expected to change), and like nearly all *de novo* non-coding mutations, these are obligate n=1 experiments. Yet, of the three antibodies that gave interpretable results, we did observe that two of these proteins appeared to have altered expression, suggesting these are *bona fide* functional mutations (*SMC5* appeared unchanged at baseline, and *C12orf57* did not have suitable antibodies to further assess).

Taking positive results from luciferase, AEI/ATI, and immunoblotting together, our *in cellulo* screen led us to the discovery of up to 19 5′UTR variants from autism cases, including five for which we have at least some evidence that they functionally altered translation of their host genes, and thus have potential to be causal for these patients. Three of these five genes (the E3 Ligase *MKRN1*, the de-ubiquitinase USP48, and the cohesin-complex-like protein SMC5) have no previous association to autism or intellectual disability from prior exome studies. However, *MKRN1* and *USP48* are highly constrained, which is a signature of many neurodevelopmental disorder genes. Likewise, the mutation we observed in *SMC5* actually is gain of function (GOF), and GOF is not as readily identified from sequencing studies alone. Thus it is unclear if *SMC5* GOFs may have recurrent consequences on human brain development, and we did not see increase in protein levels in patient LCLs by western blot at steady state. Of the remaining genes impacted by variants screened for AEI and ATI, the orphan nuclear receptor *NR4A2*, the histone methyl transferase *NSD2*, and the uncharacterized gene *C12orf57* all have some evidence for association to NDD and autism (scores of 1, 2S, and S in SFARIgene, respectively), and the first two are quite constrained in the human population. *NR4A2* in particular has been reliably associated with NDD, with nine patients carrying heterozygous missense and loss-of-function LOF mutation resulting in developmental delay (9/9), behavioral problems (5/9), language impairment (5/9) and frequent epilepsy (6/9) in a recent case series (Singh et al., 2020). Our proband for this mutation, 14147.p1, has a relatively milder phenotype without epilepsy or frank intellectual disability, though a clear diagnosis of autism. This milder presentation could be consistent with a more moderate effect of the mutation, which only decreased protein production by 15% in the reporter assay which is far from haploinsufficiency in cases of LOF mutations. Meanwhile, *NSD2* mutations cause Rauch-Steindl syndrome, which itself is a milder version of Wolfs-Hirschhorn syndrome caused by a multigene deletion encompassing this syndrome. A case series of 28 Rauch-Steindl patients revealed a consistent but mild developmental delay, frequent autistic features (33%), anxiety (15%), hyperactivity (22%), and aggressiveness (11%)(Zanoni et al., 2021). Gastrointestinal issues were also common (44%). Our proband 13654.p1 also exhibited no epilepsy, but did have autism, and higher scores for ADHD, anxiety, and oppositional behavior, along with unusual stools, which is potentially consistent with gene dose influencing these phenotypes — though he did not have the short stature or IDD reported in many stop-gain carriers. Finally, *C12orf57* mutation causes substantial brain malformations (Temtamy syndrome) when both copies are mutated (Akizu et al., 2013), but any phenotypes for heterozygous carriers are unclear.

Finally, as autism is thought to be due to changes in neurons, we developed a new approach to assess the impact of allelic variants in neurons in the developing brain via MPRA. Building on our prior work where we had shown the ability to assess the effects of 3’ UTR elements (but not at that point alleles) in specific cell types *in vivo (Lagunas et al., 2023)*. To enable a more sensitive assay, we made several adaptations here. Specifically, (1) we increased the number of barcodes per allele to improve within experiment replication, (2) decreased library complexity by breaking the elements into smaller sublibraries to insure each element was delivered to more cells, and (3) increased the expression levels by use of a CMV reporter and spliced intron. The one remaining area for future improvement might be to better optimize the assay to disentangle transcriptional from post-transcriptional effects on RNA abundance measures, as current assays would not, and thus some effects at that level could be due to allelic effects on promoter activity. However, this would not be expected to account for the majority of our allelic findings, which mostly influenced stages of ribosome occupancy rather than merely transcript abundance. Finally, we also redesigned the Cre/Lox responsive cassette to only flank the primer site, removing the requirement to have LoxP sites in the 5′UTR. These innovations enabled us to reproducibly detect allelic effects in specific cell types *in vivo*, highlighting an additional 9 variants from patients that may have neuron-specific consequences, including three in suspected autism or genes *LRFN5, LRRC4, and ZNF644.* Of these, the prior evidence for autism association of *LRFN5* was more modest and from common variant association (Wang et al., 2009). Patients with functional *LRFN5* mutations specifically have not been deeply characterized. *LRRC4* showed association in earlier exome studies (Satterstrom et al., 2020), driven more by missense variation, though the signal was a bit more modest in the most recent reanalysis (TADA-NDD FDR <0.02)(Fu et al., 2022). However, *ZNF644* shows strong association with neurodevelopmental disorder (TADA-NDD FDR <0.005), driven primarily by developmental delay rather than autism per se. Previous work also associated missense mutation of this gene with myopia(Shi et al., 2011), but neurocognitive characterization of this disorder has not been described. While from our *in cellulo* luciferase, ATI/AEI, and immunoblotting studies, we identified several case mutations for which there was further supporting evidence of functional consequences, and several more for which LCLs were not informative. Likewise, *in vivo* luciferase assays for allelic effects in neurons *in vivo* are not feasible, nor are any patient brains available for western or AEI/ATI studies. Yet, our *in cellulo* pipeline suggests a measurable subset of these *in vivo* defined variants may also alter final protein production in the brain, and thus have potential to contribute to pathogenesis.

## Methods

### 5′UTR Conservation Analysis

To analyze the conservation of 5′UTR sequences we took genes with expression measured by TPM >1 in the GTEx database (v8) (GTEx Consortium, 2020) and pulled the 5′UTR sequence of their longest transcript. These sequences were then run through both PhastCons(Pollard et al., 2010) and PhyloP (Siepel et al., 2005) pipelines using the corresponding UCSC (Kent et al., 2010) bigWig files which give conservation scores for the alignment of 99 vertebrate genomes to the human genome. To compare various groups of UTRs, we subsetted them first by their inclusion in the SFARI gene database (Abrahams et al., 2013). SFARI genes were then further grouped by their score, 1-3 with 1 being a gene clearly linked to ASD and 3 being assigned to genes with suggestive evidence of ASD implication. GTEx genes not included in the SFARI database were then run through the BioconductoR package, TissueEnrich(Jain and Tuteja, 2019). Due to unique features of UTRs in the brain, we wanted to ensure any results we were observing in the SFARI genes were in fact unique to the SFARI genes rather than neuronal genes as a whole. Thus, the TissueEnrich package allowed us to subset out those genes enriched in the brain, allowing us to compare the various SFARI genes with both brain-enriched and non-brain enriched genes. Statistical significance was determined by Kruskal-Wallis one-way ANOVA followed by a post-hoc pairwise Wilcoxon t-test.

### Library Design

We screened all *de novo* variants from both probands and unaffected sibling controls reported by (An et al., 2018) to identify those that were to occur in any annotated 5′UTR using the utr.annotation R package (Liu and Dougherty, 2021). For each variant, all Ensembl (v99) annotated transcript isoforms were considered to identify all non-degenerate sequence contexts. Within each context, a maximum of 220 nt was taken, centering the position of variation when possible. If the variant occurred near the annotated 5′-end of a transcript isoform, the UTR sequence was extended to include additional sequence at the 3′-end up to 220 nt total. For variants found within 110 nt of the annotated start codon, the 3′-end of the variant context was trimmed to include no more than the first six codons of the coding sequence. The enumerated transcript contexts for each variant were de-duplicated, retaining all unique sequence contexts for each allele of the variant. From this set of potential 5′UTR sequences, we removed any which contained restriction sites for AgeI, KpnI, BamHI, and SalI. For cloning, AgeI and BamHI restriction sites were added to flank the 5′UTR sequence. For 5′UTR sequences shorter than 220 nt, padding sequence was added after the BamHI site. After the BamHI site or padding sequence, a KpnI site was added followed by a 10 nt barcode sequence followed by SalI restriction site. Primer binding sites 20 nt in length were appended to the ends of each sequence. To allow modularity in library cloning and viral packaging, the 5′UTR variant sequences were binned into three separate libraries of approximately 500 variants each using distinct primer sets to facilitate targeted amplification. Each variant-derived 5′UTR sequence was barcoded with 10 unique sequences. A fourth library subset was designed with its own unique primer set and contained a set of controls. A “blank” 5′UTR control containing no insert between the AgeI and BamHI sites was included with 100 unique barcodes. A set of 300 randomized shuffled sequences were generated to sample the range of GC content spanned by the variant library, each with 10 unique barcodes. Last, a set of natural and synthetic controls were included with representative 5′UTR elements, including uORFs, uAUGs, hairpins, and G-quadruplexs. Each of these representative control elements was barcoded 50 times. See **Supplemental Table 1** for complete details. In total, the library consisted of 1,507 allelic pairs of reporters from 997 variants (526 from probands and 471 from unaffected siblings). With controls, the complete library consisted of 32,990 uniquely barcoded elements.

### Library Cloning

The designed 5′UTR-BC library was synthesized via oligo array synthesis by Twist Bioscience. ssDNA oligos were amplified with 12 cycles using NEBNext High-Fidelity 2x PCR Master Mix using one of four primer sets to enrich a specific library subset (primers in **Supplemental Table 6**). Following PCR column cleanup, the PCR products were digested with BamHI and SalI, size selected with a 1.8:1 SPRI bead:sample cleanup, and ligated into pJD460 which comprised the following features: flanking ITRs for AAV packaging, a CMV promoter 32 bp upstream of the BamHI restrict site, and an hGH terminator. Ligation products were electroporated into competent cells prepared in-house from NEB Stable chemical competent cells (NEB C3040). A Maxi-prep (Qiagen) of the transformed first-step library was prepared and digested with AgeI and KpnI, into which a tdTomato coding sequence modified with an N-terminal Myc tag and chimeric intron was ligated. The second-step ligation was again electroporated into NEB Stable cells, and the final plasmid library was obtained by Maxi-prep. Representation of all barcodes in the library was confirmed by high-throughput sequencing as described below.

### Cell Culture and Transfection

HEK cells were submitted for STR profiling by ATCC and matched ATCC HEK293 cells with 81% similarity in tested markers. These cells were found to be negative for mycoplasma by the Genome Engineering and Stem Cell Center at the McDonnell Genome Institute. Cells were maintained at 5% CO2, 37°C, 95% relative humidity in 1:1 Dulbecco Modified Eagle Medium/Nutrient Mixture F-12 (DMEM/F12, GIBCO) supplemented with 10% fetal bovine serum (FBS, Atlanta Biologicals) and 1% Penicillin-streptomycin (Gibco). Cells were passaged every 2-3 days or once 80% confluent using 0.25% Trypsin-EDTA (Gibco).

For transient transfection of the MPRA plasmid library, 6.75 million cells were seeded per T75 flask 24 hours prior to transfection in DMEM/F12 supplemented with 10% FBS. Each replicate flask was transfected with 18 ug of the MPRA library plasmid mixed with 2 ug of pCMV-eGFP-Rpl10a using Lipofectamine 2000 (Invitrogen). Media was changed 6 hours following transfection, and cells were collected 48 hours post-transfection.

### RNA & DNA isolation from cultured cells

To harvest RNA for polysome fractionation, cultured cells were lifted into the plating medium and pelleted by centrifugation at 300x*g* for 3 minutes. Cell pellets were resuspended in ice-cold PBS with 100 µg/mL cycloheximide and briefly kept on ice. Cells were re-pelleted by centrifugation at 500x*g* for 3 minutes and resuspended in 1mL of Lysis Buffer (20 mM HEPES, 150 mM KCl, 10 mM MgCl2, 0.5 mM DTT, 1% NP-40, 100 µg/mL cycloheximide, 0.02 U/µL rRNasin, 0.02 U/µL Superasin, EDTA-free protease inhibitor at 1 tablet per 50 mL). Cells were triturated with ten passes through a 27 gauge needle. Lysates were clarified by centrifugation at 20,000x*g* for 10 minutes at 4°C. To collect total RNA, 50 µL of the clarified lysate was taken for extraction using Trizol LS (Invitrogen) reagent followed by treatment with DNase I (Zymo) and column clean-up (Zymo). Precipitated DNA taken from the Trizol interphase was resuspended in PBS and re-concentrated using the Qiagen DNeasy Blood & Tissue kit. The remaining clarified lysate was split into two fractions (350 µL for polysome fractionation and 600 µL for TRAP) and snap frozen in liquid nitrogen.

### Translating Ribosome Affinity Purification (TRAP)

TRAP was performed as described previously(Heiman et al., 2008) with minimal modifications. Snap frozen lysates from HEK cells were thawed on ice before adding DHPC (Avanti) to 30 mM final concentration. Lysates were further incubated before spinning at 20,000xg for 10 minutes at 4°C. Anti-eGFP magnetic beads were prepared by overnight incubation of streptavidin-coated MyOne T1 Beads (Invitrogen) with biotinylated Protein L (Thermo Scientific) and two monoclonal anti-eGFP IgG antibodies (clones 19F7 and 19C8)(Doyle et al., 2008). The final clarified suerpantant was incubated with the anti-eGFP beads for 6 hours at 4°C followed by four washes with 1mL High Salt Buffer (20 mM HEPES, 350 mM KCl, 10 mM MgCl2, 0.5 mM DTT, 1% NP-40, 100 µg/mL cycloheximide, 0.02 U/µL rRNasin, 0.02 U/µL Superasin, EDTA-free protease inhibitor at 1 tablet per 50 mL). After the final wash, the beads were resuspended in 250 µL Lysis Buffer and added to 750 µL Trizol LS. RNA was extracted from Trizol followed by DNase I treatment and column clean-up.

### Polysome fractionation

Sucrose gradients were prepared by layering 2 mL of buffered sucrose solutions (20 mM HEPES, 150 mM KCl, 10 mM MgCl2, 1 mM DTT, 100 µg/mL Cycloheximide) ranging from 50 to 10%, freezing at -80°C between the addition of subsequent layers. Gradients were thawed and allowed to linearize overnight at 4°C. Lysates were layered over thawed gradients and spun at 151,000 g for 2.5 hours at 4°C using a Beckman SW-41 Ti rotor. Fractions of 500 uL were collected by upward displacement of the gradients using a Teledyne ISCO fractionator while monitoring absorbance at 254 nm. Fractions corresponding to 40S, 80S, and polysomes were pooled separately. RNA from the 40S and 80S fractions was extracted using Trizol LS, followed by treatment with DNase I (Zymo) and column clean-up (Zymo). RNA from polysome containing fractions was precipitated by the addition of 0.1 volumes of 4 M Sodium Acetate pH 5.2 and 3 volumes of ethanol and freezing overnight at -20°C. Pelleted polysome RNA was treated with DNase I and concentrated by column clean-up similar to 40S and 80S RNA.

### AAV9 vector production

The packaging cell line, HEK293, is maintained in Dulbecco’s modified Eagles medium (DMEM), supplemented with 5% fetal bovine serum (FBS), 100 units/ml penicillin, 100 mg/ml streptomycin in 37°C incubator with 5% CO2. The cells are plated at 30-40% confluence in CellSTACS (Corning, Tewksbury, MA) 24 h before transfection (70-80% confluence when transfection). 960 ug total DNA (286 ug of pAAV2/9, 448 ug of pHelper, 226 ug of AAV transfer plasmid) are transfected into HEK293 cells using polyethylenimine (PEI)-based method with modifications.(Challis et al., 2019) The cells are incubated at 37°C for 3 days before harvesting. The cells are lysed by three freeze/thaw cycles. The cell lysate is treated with 25 U/ml of Benzonaze at 37°C for 30 min and then purified by iodixanol gradient centrifugation. The eluate is washed 3 times with PBS containing 5% Sorbitol and concentrated with Vivaspin 20 100K concentrator (Sartorius Stedim, Bohemia, NY). Vector titer is determined by qPCR with primers and labeled probe targeting the ITR sequence(Aurnhammer et al., 2012).

### Animal Models

Veterinary care and housing is provided by the veterinarians and veterinary technicians of Washington University School of Medicine and all procedures were approved by the Institutional Animal Care and Use Committee. The colony room lighting was a 12–12 h light–dark cycle, and room temperature (∼20–22 °C) and relative humidity (50%) were controlled automatically. Standard laboratory diet and water were freely available. All protocols involving animals were completed with: Ef1a:LSL-eGFP-RPl10 (Jackson Stock no: 030305) and Vglut1-IRES2-Cre-D strain (Jackson Stock No: 023527). All mice were genotyped following a standard protocol of taking clipped toes into lysis buffer (0.5M Tris-HCl pH 8.8, 0.25M EDTA, 0.5% Tween-20, 4uL/mL of 600 U/mL Proteinase K enzyme) for 1 hour to overnight. This is followed by heat denaturation at 99 C for 10 minutes. 1 uL of the resulting lysate was used as a template for PCR with 500 nM forward and reverse primers, using 1x Quickload Taq Mastermix (NEB) with the following cycling conditions: 94 1 min, (94 30 sec, 60 30 sec, 68 30 sec) x 30 cycles, 68 5 min, 10 hold.

### *in vivo* MPRA

Pups were genotyped at day P0, and those positive for eGFP and Cre were transduced at day P1 with a mixture of two AAV9-packed libraries: a pool of variant reporters consisting of approximately 500 allelic pairs (about 10,000 total barcodes) and a pool of control reporters consisting of 2,850 barcodes. For transcranial delivery, pups were anesthetized by inducing hypothermia on ice. A 33G Hamilton syringe was used to inject 1 µL of the AAV preparation (titer 10^13^ viral genomes/µL) at three positions per hemisphere (6 total injections per pup). Pups were recovered on a warming pad before placing back into the cage with the mother and monitored every 24 hours for one week.

At day P21, the cortex was dissected away from the striatum and washed in ice-cold homogenization buffer. The tissue was placed in a dounce tissue grinder (Kimble Kontes Catalog Number 885300-0002) in 2 mL of Lysis Buffer and homogenized with 15 strokes of the size A pestle and 15 strokes of the size B pestle. Lysates were clarified by centrifugation at 2,000xg and 4°C for 10 minutes. The supernatant was transferred to a new tube, and the remaining pellet was resuspended in 1.75 mL Trizol LS. To the clarified supernatant, NP-40 (Sigma) was added to a volume concentration of 1% and DPHC (Avanti) to a concentration of 30 mM. Supernatants were incubated on ice for 10 minutes before centrifugation at 20,000xg at 4°C for 10 minutes. A 250 µL fraction of the final lysate was collected and added to 750 µL Trizol LS while the remaining 500 µL was snap-frozen in liquid nitrogen and saved for polysome fractionation. From the 2,000xg spin pellet, DNA was extracted from the interphase of the Trizol LS phenol-chloroform extraction, according to the kit instructions. Input RNA was extracted from the clarified lysate added to Trizol LS after the 20,000xg spin.

### Fluorescence Immunohistochemistry

Whole brains from animals transduced with the AAV9-packaged reporter libraries were harvested at day P21 and postfixed in 4% paraformaldehyde for 24 h followed by 48 h in 30% sucrose 1x PBS. Brains were then frozen in OCT compound (optimum cutting temperature compound; catalog #23-730-571, Thermo Fisher Scientific). Coronal sections were taken at 40 μm thickness on a Leica CM1950 cryostat and stored in 1X PBS and 0.1% w/v sodium azide. Sections were incubated in a blocking solution (1× PBS, 5% donkey serum, 0.25% Triton-X 100) for 1 h in a 12 well plate at room temperature and then stained overnight at 4°C with rabbit anti-RFP primary antibody (1:500; Rockland catalog no. 600-401-379) and chicken anti-eGFP primary antibody (1:1000, Aves Labs catalog no. GFP-1020). The sections were washed three times for five minutes before staining with donkey anti-rabbit Alexa Fluor 568 secondary antibody (1:1000, Invitrogen catalog no. A10042) and Donkey anti-chicken Alexa Fluor 488 (1:1000, Jackson Immuno catalog no. 703-545-155). Sections were washed three times for 5 minutes, adding 1 μg/mL DAPI on the second wash. Sections were slide mounted with Prolong Gold antifade. Images were taken on a Zeiss Axio Imager Z2 four-color inverted confocal microscope at 40x magnification.

### Sequencing Library Preparation

RNA was reverse transcribed using the SuperScript IV Reverse Transcriptase kit. For fractions collected from HEK cells, RNA was reverse transcribed using the CreOFF_RT_UMI primer (IDT) which appended a 12 bp unique molecular identify (UMI) to each cDNA molecule. For RNA fractions collected from cortical RNA, the CreON_RT_UMI primer was used for reverse transcription to selectively capture mRNA from Cre-expressing Vglut2-positive neurons. The cDNA was PCR amplified with NEBNext HF 2x Master Mix (NEB) using FivePrime_Sense and FivePrime_Antisense primers. An indexing PCR was performed using NEB Ultra II Q5 2x Master Mix (NEB) and custom Illumina TruSeq adapters. DNA libraries were prepared by amplifying extracted plasmid using with NEBNext HF 2x Master Mix (NEB) using CreOFF_RT_UMI and FivePrime_Antisense primers for the HEK assay and CreON_RT_UMI and FivePrime_Antisense primers for the neuronal assay. The same PCR protocol was used to amplify barcodes from the Maxi-prepped plasmid library, and the same indexing PCR was used for DNA libraries as for RNA. Indexed libraries were quantified using HSD1000 Tapestation ScreenTapes (Agilent) and pooled for sequencing. The final libraries were sequenced by 2x151 paired-end sequencing on the Illumina NovaSeq platform by the Genome Technology Access Center at the McDonnell Genome Institute.

### Sequencing Library Analysis

UMI sequences were extracted from the start of Read2 fastq files using umi_tools (v1.0.0)(Smith et al., 2017) using the following command and arguments:

umi_tools extract -I <READ2 fastq> --bc-pattern=NNNNNNNNNNNN --read2-in=<READ1 fastq> --stdout=<PROCESSED Read2 fastq> --read2-out=<PROCESSED Read1 fastq>

Barcode sequences were extracted from the umi_tools processed Read1 and Read2 fastq files using *cutadapt* (v2.1)(Martin, 2011). Barcodes were trimmed from libraries primed with the CreOFF_RT_UMI primer with the following commanded and arguments:

cutadapt --cores=0 -a

“^TAGTAACACGACGCCGACTGTCGAC…GGTACCCGCATCGTTAATCGGCATAACTTCGTATAATGTATGCTATACGAAGTTATGGGTCGATGGTGAGATCTGGACTAGAGGGTCGANNNNNNNNNNNNAGATCGGAAGAGCAC$” -A “^TCGACCCTCTAGTCCAGATCTCACCATCGACCCATAACTTCGTATAGCATACATTATACGAAGTTATGCCGATTAACGATGCGGGTACC…GTCGACAGTCGGCGTCGTGTTACTAAGATCGGAAGAGCGT$” -o <TRIMMED Read1fastq> -p <TRIMMED Read2 fastq> --pair-filter=any --discard-untrimmed <PROCESSED Read1 fastq> <PROCESSED Read2 fastq>

Barcodes were trimmed from libraries primed with the CreON_RT_UMI primer with the following commanded and arguments:

cutadapt --cores=0 -a “^TAGTAACACGACGCCGACTGTCGAC…GGTACCCGCATCGTTAATCGGCATAACTTCGTATAATGTATGCTATACGAAGTTATCATTGGTCACGCACGATTTCNNNNNNNNNNNNAGATCGGAAGAGCACACGTCTGAACTCC$” -A “^GAAATCGTGCGTGACCAATGATAACTTCGTATAGCATACATTATACGAAGTTATGCCGATTAACGATGCGGGTACC…GTCGACAGTCGGCGTCGTGTTACTAAGATCGGAAGAGCGTCGTGTAGGGAAAG$” -o <TRIMMED Read1 fastq> -p <TRIMMED Read2 fastq> --pair-filter=any --discard-untrimmed <PROCESSED Read1 fastq> <PROCESSED Read2 fastq>

The extracted barcode sequences were aligned to an index of barcode sequences using *bowtie2* (v2.3.5)(Langmead and Salzberg, 2012):

bowtie2 --very-sensitive -p 8 --no-mixed -x <BARCODE index> -1 <TRIMMED Read1 fastq> -2 <TRIMMED Read2 fastq> -S <ALIGNMENT SAM file>

Barcode alignments were sorted using *samtools* (v1.9)(Li et al., 2009):

samtools view -bS <ALIGNMENT SAM file> > <ALIGNMENT BAM file>; samtools sort <ALIGNMENT BAM file> > <SORTED BAM file>

Reads per barcode were counted using *featureCounts* (*subread* v2.0.0)(Liao et al., 2014):

featureCounts -p -B -P -d 10 -D 10 -T 4 -t BC -g barcode -a <.gtf file with barcodes> -R BAM <SORTED BAM file> -o <FEATURECOUNTS outfile>

The alignment output by *featureCounts* was resorted and indexed:

samtools sort <FEATURECOUNTS sorted BAM file> > <RESORTED BAM file>; samtools index <RESORTED BAM file>

Finally, reads were de-duplicated to obtain UMI counts per barcode using *umi_tools*:

umi_tools count --paired --per-gene --gene-tag=XT -I <RESORTED BAM file> -S<UMI counts per barcode>

Barcode UMI counts were used in all analyses.

### MPRA Variant Effects Analysis

To test for an effect of allele on transcript abundance or measures of translation, barcodes were first filtered to remove any which had less than 20 UMI counts within a replicate for either fraction of a particular ratiometric measurement. Variants were not tested for allelic effects if they had fewer than three biological replicates with at least three barcodes for reference and alternative each after filtering. For testing for effects of Cre and allele-Cre interactions, CreON and CreOFF libraries had to share at least three replicates with three or more ref and alt barcodes. After filtering, the log2-ratio of counts between fractions was computed for each barcode within each biological replicate, and all barcode log2-ratios were scaled relative to the mean value of the 100x barcoded blank control. To test for allelic effects, the log2-ratios were fit to a linear mixed effect model using the *lme4(Bates et al., 2015)* package in R, including a random intercept term for barcode: logratio ∼ Allele + (1|BC). Null comparisons were sampled from the set of blank barcodes, randomly assigning 10 barcodes as “reference” and 10 as “alternative” and repeating a total of 10,000 times. The distribution of test statistics from the allele term of these null comparisons was used to compute an empirical p-value for each of the variants which were then used to calculate multiple test corrected q-values using the *qvalue* package (Storey et al., 2023). Allelic comparisons with q-value < 0.05 were considered significant changes. For *in vivo* experiments, each sublibrary of variants was analyzed for allelic effects separately within each context (CreON or CreOFF) except when testing for the effect of Cre or allele-Cre interactions. These effects were evaluated with the interaction model: logratio ∼ Allele + Cre + Allele:Cre. Empirical p-values and q-values were computed for each model term with the same procedure used for the allele-only models.

### uORF feature analysis

The start codon context for uORFs was categorized as weak, medium, or strong based on pattern matching (strong: (A|G)NN<u>AUG</u>G; medium: NNN<u>AUG</u>G or (A|G)NN<u>AUG</u>Nl weak: NNN<u>AUG</u>N) (Whiffin et al., 2020). When evaluating the effect of uORF length or the distance between uORF stop codons and reporter gene start codons, the first uORF in a 5′UTR that had medium or strong uAUG context was considered.

### RNA structure prediction

The minimum free energy of folding was calculated for the reference and alternative allele of each allelic reporter pair using the ViennaRNA package (v2.4.17) (Lorenz et al., 2011). Predicted folding structures for the *SMC5* reporter alleles were generated using the RNAfold webserver (Gruber et al., 2008). G-quadruplex structure for the *SMC5* reporter alleles was predicted using QGRS Mapper (Kikin et al., 2006).

### DNA and RNA binding motifs

Changes in the number of DNA and RNA binding motifs as a result of each variant were assessed by scanning the reference and alternative 5′UTR sequences with sets of position-weighted matrices (PWMs) using code and motif sets adapted from Lim et al. 2021(Lim et al., 2021). DNA-binding element PWMs were previously generated for a set of 332 motifs obtained from the HOMER database(Weirauch et al., 2014), and 102 RBP motifs were obtained from (Ray et al., 2013). Motif occurrences were scored using a custom Python script which scanned PWMs over each reporter 5′UTR sequence, counting every k-mer with a score greater than or equal to 90% of the maximum possible score for each motif. DNA-binding motifs were scanned over the sense and antisense strands; RBP motifs were only scanned over the sense strand. For each set of motifs, one-sided Mann-Whitney U tests were performed to determine whether variants which altered the number of motifs (either as a gain or loss) had on average greater absolute effect sizes on transcript abundance or translational measures compared to the set of variants which had no effect on motif occurrence.

### Dual-Luciferase Reporter Assay

HEK cells were plated at a density of 2.5 x 10^4^ cells per well in white-walled, clear-bottom 96-well plates (Corning) coated with 0.01% poly-*L*-lysine (Sigma). The cells were cultured for 24 hours prior to transfection in DMEM supplemented with 10% FBS. The cells were transfected with a mixture of 180 ng of a pCMV-eGFP carrier plasmid and 20 ng of a dual luciferase reporter plasmid using Lipofectamine 2000 (Invitrogen). Each dual luciferase plasmid included a 5′UTR of interest cloned ahead of a NanoLuc-PEST ORF driven by a CMV promoter and firefly luciferase ORF driven by a PGK promoter. The media was replaced 6 hours after transfection with DMEM 10% and 1% penicillin-streptomycin which did not contain phenol red. Transfected cells were incubated for an additional 17 hrs. Luminescence was finally read using Nano-Glo Dual-Luciferase Reporter Assay System on a Cytation 5 multi-modal plate reader. Four replicate wells were used for each allele, and variant pairs were run together on the same plate. Luminescence values for both NanoLuc and firefly luciferase were blank subtracted using the signal from wells transfected only with the carrier plasmid. The resulting NanoLuc luminescence was normalized to the firefly luminescence in each well. A control reporter plasmid that did not contain a 5′UTR insert was run on each plate, and the luminescence ratio for the control plasmid was used to normalize the luminescence ratio of each well on the same plate. Control-normalized luminescence log2-ratios of NanoLuc and firefly luminescence were correlated against MPRA measures transcript abundance, 80S/40S enrichment, polysome/80S enrichment, and polysome/total RNA enrichment. Mann-Whiteny U tests were used to test for a difference in the log2 luminescence ratio between reference and alternative alleles and corrected for multiple tests using a Benjamini-Hochber FDR.

### Allelic Expression and Translation Imbalance Analyses in Patient Lymphoblasts

Patient-derived lymphoblast lines were obtained from SFARI Base. Upon thawing, cultures were maintained in Roswell Park Memorial Institute (RPMI) 1640 supplemented with 15% FBS and 1% penicillin-streptomycin. After 72 hours in culture, cells were harvested for polysome fractionation as described above. Fractionated RNA was reversed transcribed using the SuperScript III Reverse Transcriptase kit and a gene specific primer (**Supplemental Table 6**). The cDNA was PCR amplified with NEB Ultra II Q5 2x Master Mix (NEB) using gene specific primers. An indexing PCR was performed using NEB Ultra II Q5 2x Master Mix (NEB) and custom Illumina TruSeq adapters. DNA libraries were prepared by amplifying extracted genomic DNA with NEB Ultra II Q5 2x Master Mix (NEB) using gene specific primers. The same indexing PCR was used for DNA libraries as for RNA. The final libraries were sequenced by 2x150 paired-end sequencing on the Illumina MiSeq platform by the DNA Sequencing Innovation Lab at the Center for Genome Sciences and Systems Biology.

From the sequencing reads, the reference or alternative allele for each target were counted using a custom Python script. To asses AEI and ATI, negative binomial generalized linear models(Robinson et al., 2010) were fit to determine if the alternative allele was differentially abundant across two fractions (total RNA / DNA for AEI; 80S / 40S, polysome / 80S, or polysome / total RNA for ATI). For each variant, dispersion was estimated for each allele across all sequenced libraries.

### Western Blotting

Commercially available antibodies for MKRN1, NSD2, SMC5, C12orf57, and TPSAN4 were tested by using them to blot in lysates from LCLs of patients and siblings obtained from SFARIbase. Of these, only blots with antibodies for MKRN1, NSD2, and SMC5 detected bands of the correct predicted size for the target protein.

Once pelleted from cell culture, cells were washed in ice-cold 1x PBS and centrifuged at 2,000 g for 7 min at 4C. Supernatant was extracted and discarded. Lysis buffer consisting of 10x RIPA Buffer (Cell Signaling, 9806) diluted to 1x with water and 50x proteinase inhibitor diluted to 1x was added to pelleted cells. Cells were agitated for 30 minutes at 4C. Samples were then centrifuged at 16,000 g for 20 min at 4C and supernatant collected. Protein concentration was determined using PierceTM BCA Protein Assay Kit (Thermo Scientific, 23227). Samples were boiled with 2x Laemmli Sample buffer for 5 min and 50µgs were loaded onto Mini-Protean TGX gels (Bio-Rad, 456104). Gels were electrophoresed at 80V for 10 minutes, and then 150V for 45 minutes. After, gels were washed in transfer buffer (1x SDS-PAGE Running Buffer without SDS, 20% methanol) twice for 10 minutes each. Immun-Blot PVDF membrane (Bio-Rad, 1620177) was activated in 100% methanol, washed for 10 minutes in deionized water, and equilibrated in transfer buffer for 10 minutes. Protein from gels were transferred to membrane using Trans-blot SD Semi-Dry transfer cell (Bio-Rad) using 2*area of gel (cm)/1 hour to determine mAmps. After transfer, the membrane was washed in 1X TBST 2 times for 5 minutes each. The membrane was then blocked for 1 hour at room temperature in 1X TBST + 5% Milk. Primary and secondary antibodies were diluted in block. Primary antibodies used were rabbit anti-MKRN1 1:1500 (abcam, 72054), mouse anti-SMC5 1:100 (Santa Cruz, sc-393282), mouse anti-NSD2, clone29D1 0.5ug/mL (Milipore Sigma, MABE191), mouse anti-GAPDH 1:10,000 (Sigma-Aldrich, G8795). Secondary antibodies were 1:2000 rabbit-HRP and1:2000 mouse-HRP. The membrane was incubated in primary antibody block overnight in 4C, then washed in 1X TBST three times for 5 minutes. The membrane was incubated in secondary antibody block for one hour at room temperature and washed as before. After washes, the membrane was incubated in a 1:1 mixture of peroxide solution and luminol/enhancer solution (ClarityTM Western ECL Substrate, Bio-Rad) and imaged using MyECL Imager (Thermo Scientific).

## Data and Code Availability

All sequencing data, including raw fastq files and processed count files, were deposited to the Gene Expression Omnibus (GEO) database at accession number GSE246381. Unless otherwise stated all analyses and plot creation were performed in R version 4.3.0 with RStudio version 2023.09.0+463. Code and other software used to perform analyses were uploaded to BitBucket at https://bitbucket.org/jdlabteam/fiveprime_mpra_pub/src/main/. For access, please see and follow the directions outlined in the README page. Cartoon depictions of experiments/constructs (Figs 1A-B, 4B, 5A-B, and 6) were created with Biorender.com. Other resources and datasets used are appropriately cited throughout the text.

## Acknowledgments

We’d like to thank Jess Hoisington-Lopez and MariaLynn Crosby of the CGS for assistance with sequencing, Mingjie Li of the Hope Center, GTAC@MGI and GESC@MGI for their support, and finally members of the Autism Sequencing Consortium and the Dougherty lab for helpful discussions and advice. Funding was provided by SFARI 571009 and NIMH R01MH116999 to JDD, IDDRC P50HD103525, R01GM11282 to SD and R01GM136823 to SD and SPD.

---

**Figure S1:**
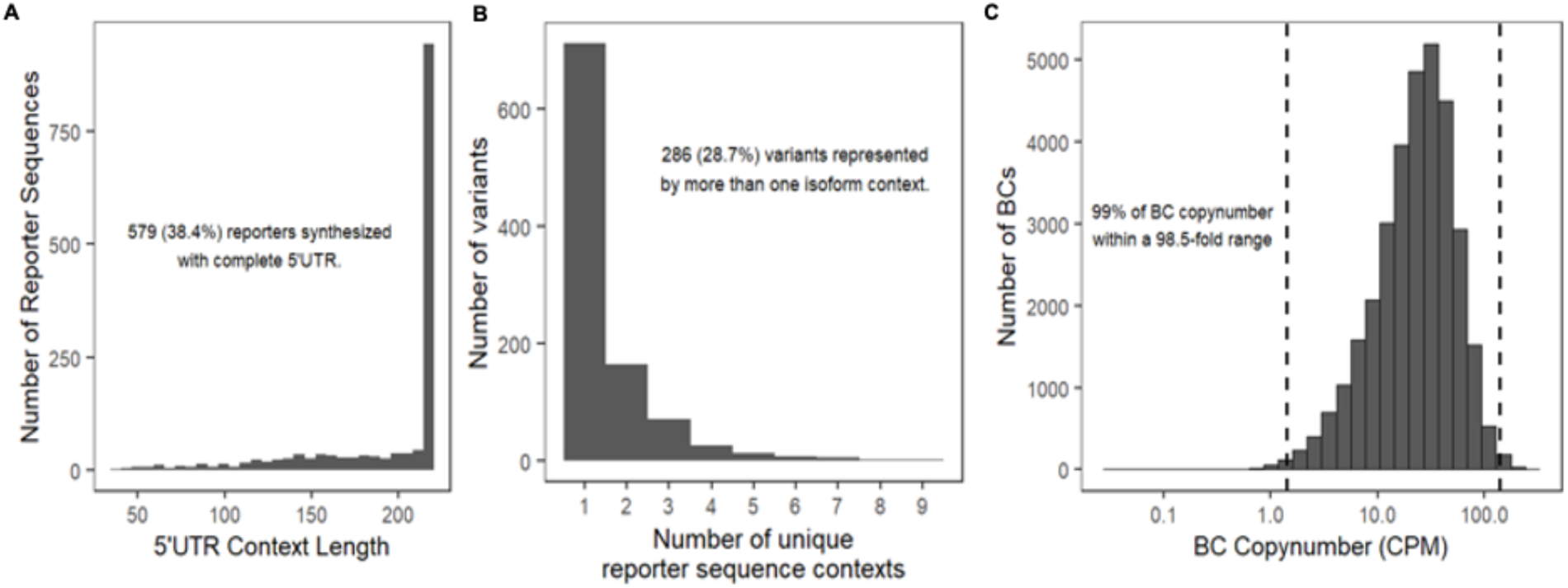
Library design statistics and barcode copy number distribution. **A)** Distribution of sizes of 997 annotated 5’UTRs for those carrying mutations in the cohort: 38.4% of reporters included the full annotated 5′UTR sequence for the annotated transcript isoform. **B)** Every Ensembl-annotated transcript isoform that spanned the position of variation was considered for each variant, and all unique sequence contexts were included in the library, resulting in 1507 sequences. **B)** Following cloning, the abundance distribution for all 32,990 barcoded reporters spanned a 100-fold range.

**Figure S2:**
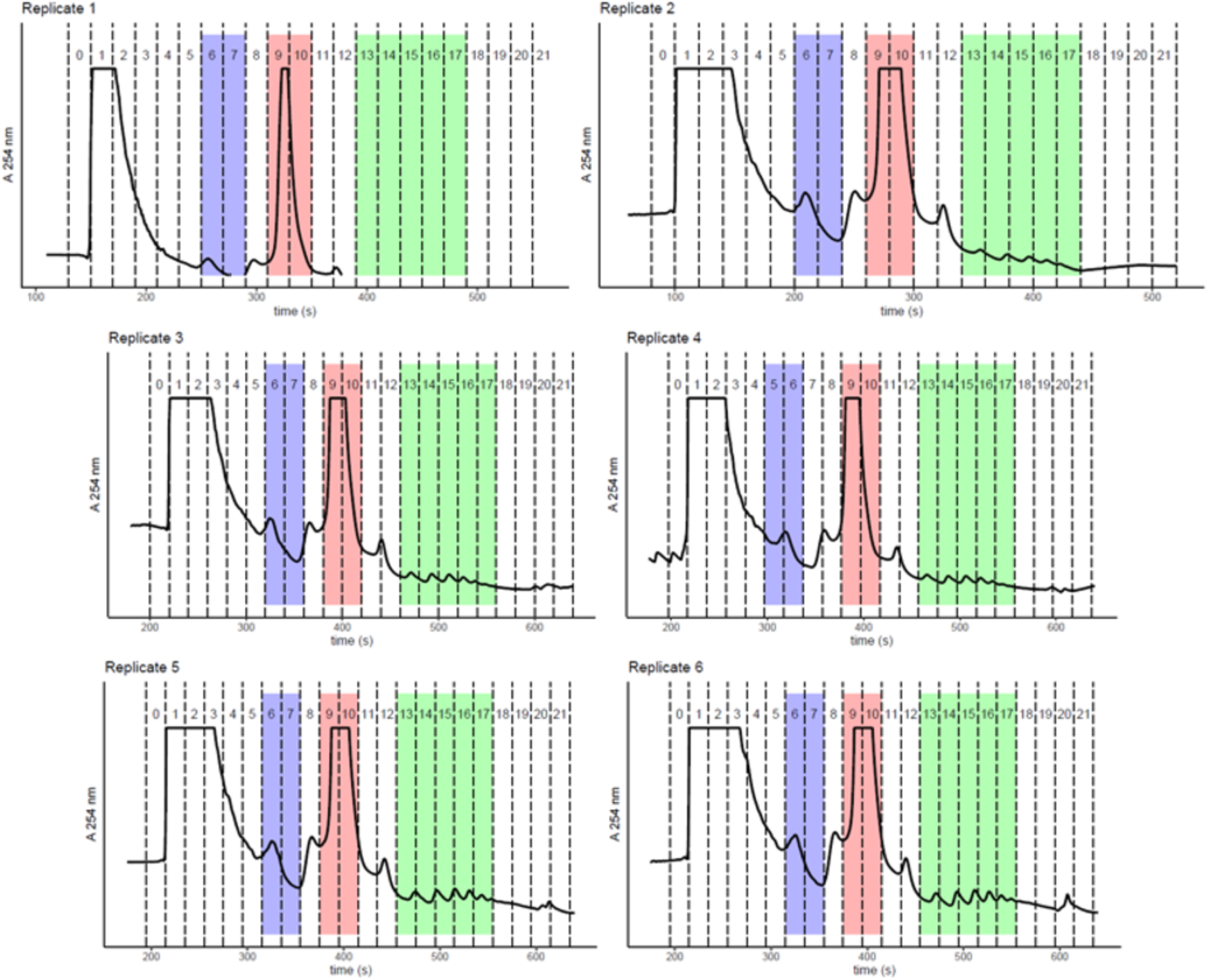
Polysome fraction traces for HEK MPRA cell lysates. Lysates from HEK cells transfected with the MPRA library were fractionated by using 10-50% sucrose gradients. Fractions were collected by upward displacement, and absorbance at 254 nm was monitored. 40S-associated RNA was extracted from fractions 6 and 7. Monosome-associated RNA was collected from fractions 9 and 10. Polysome-associated RNA was collected from fractions 13 through 17. Polysome peaks in replicate 1 were below the detector baseline, but clear polysome peaks are visible in subsequent samples following baseline readjustment.

**Figure S3:**
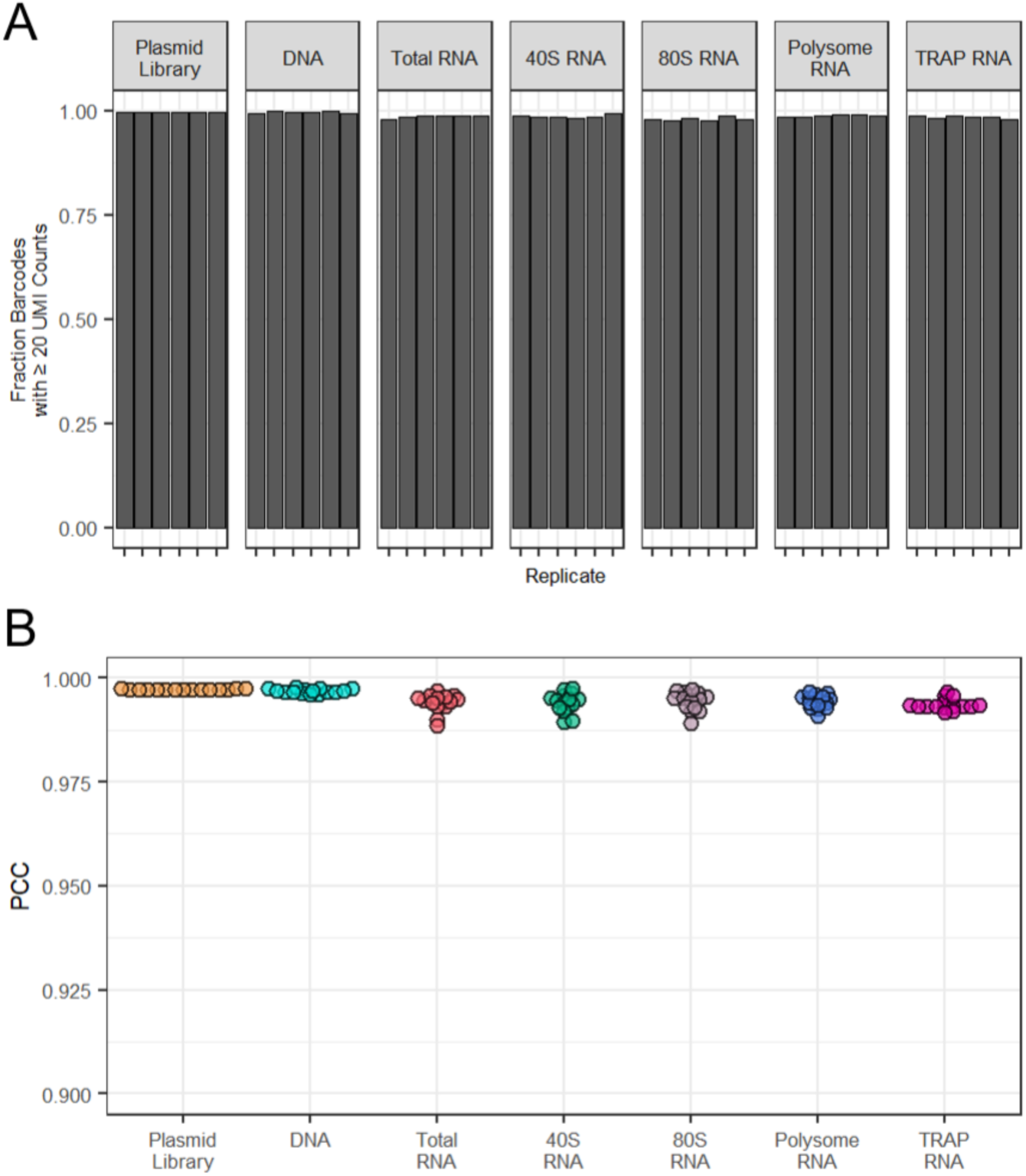
Barcode recovery rate and pairwise replicate correlations from HEK MPRA libraries. **A)** Across all fraction types, >97% of barcodes were recovered with 20 or more UMI counts. **B)** All pairwise comparisons within each fraction type show excellent correlation between replicate transfections of the MPRA library into HEK cells (Pearson’s correlation coefficient, PCC ≥ 0.988 for all comparisons).

**Figure S4:**
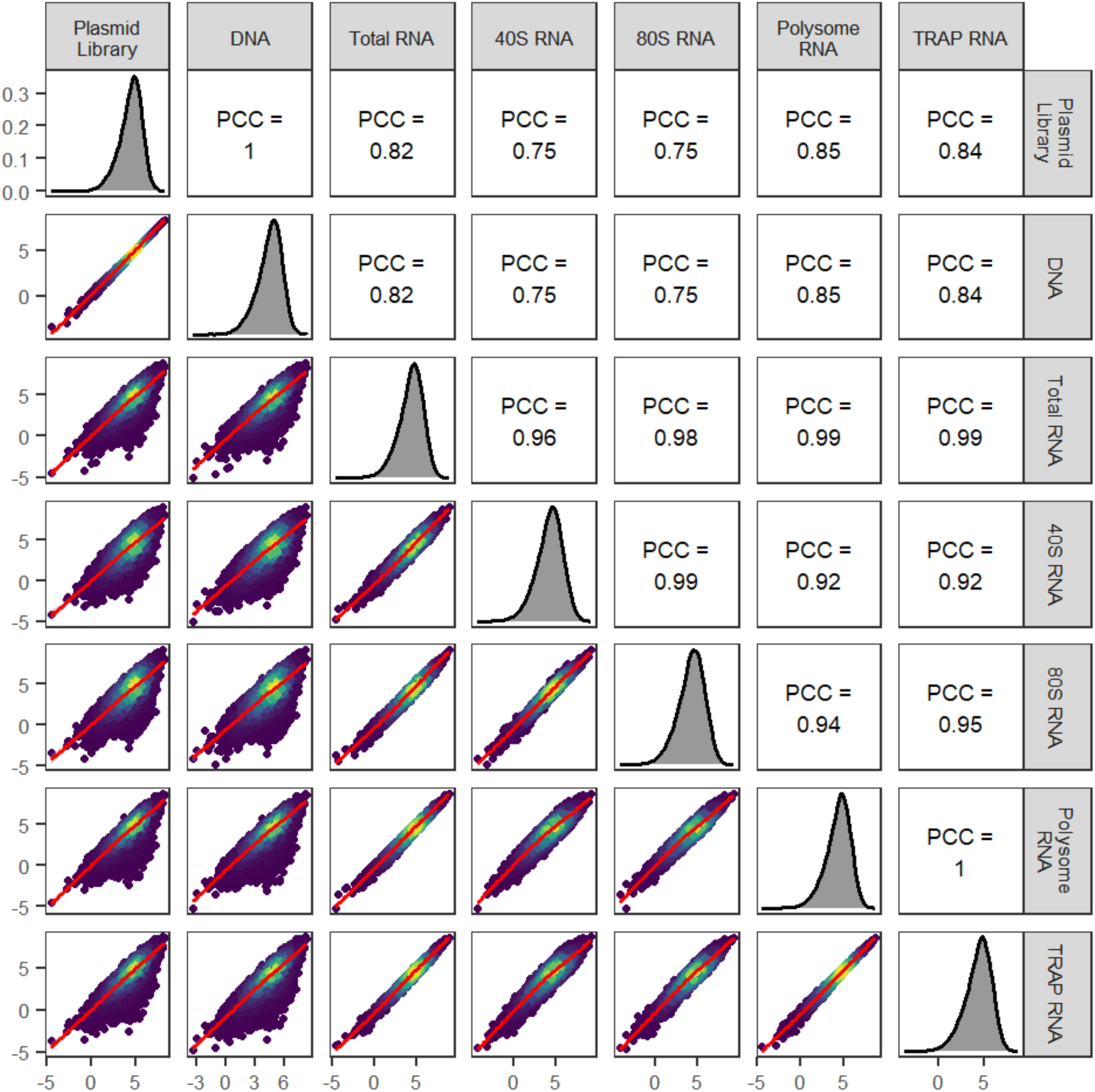
Correlations of barcode abundances between library types from the HEK MPRA. Barcode abundance measurements (plotted as log2-transformed averages across replicates) are less well correlated between RNA and DNA fractions than within the set of either RNA or DNA libraries, reflecting the biological effect on the RNA. Notably, barcode abundance in polysome-associated RNA correlates most strongly with TRAP RNA abundance, indicating TRAP-IP enriches for polysome-bound RNA over monosomes as expected. Counts from plasmid DNA recovered from lysed cells also closely correlate with the abundance in the library pre-transfection.

**Figure S5:**
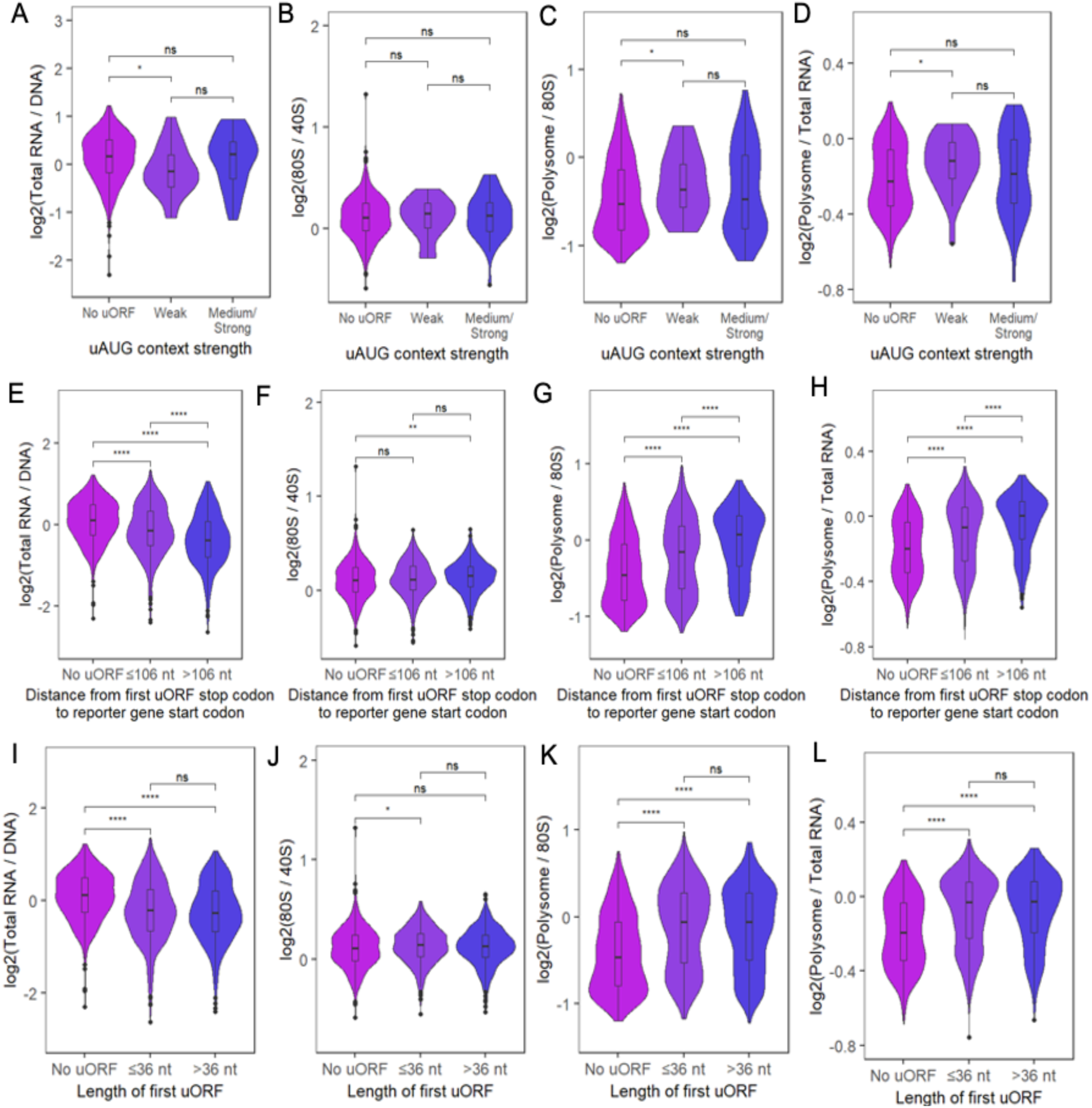
The effect uORFs have on transcript abundance and translation depends on the context of uORF start codons and the position of uORF stop codons within 5′UTR. **A-D)** The effect of uORF start codon context strength was evaluated within a set of reference allele reporters which contained either no uORF or a single uORF. Start codon strength was scored using the scheme from ref(Whiffin et al., 2020) (plotting 521 without uORFs, 24 with weak uAUG context, and 46 with either medium or strong uAUG context). Nominally significant effects on transcript abundance and certain translation measures were observed for uORFs with weak start codon contexts only. **E-H)** The impact of uORF stop codon distance from coding region start codon was examined, finding that uORFs terminating further from the reporter coding sequence had larger effects on transcript abundance and translation. The analysis used only the ‘reference allele’ reporters, and only the first uORF with a medium or strong uAUG context was considered. These reporters were split into bins by the median length between uORF stop codons and the reporter gene (106 nt). **I-L)** The same set of reporters was used to test for the impact of uORF length on transcriptional and translational effects, but no effects were found on any measure. Reporters were split into bins by the median length of the uORFs (36 nt). (Mann-Whitney U test: * p<0.05, ** p < 0.01, **** p < 0.0001)

**Figure S6:**
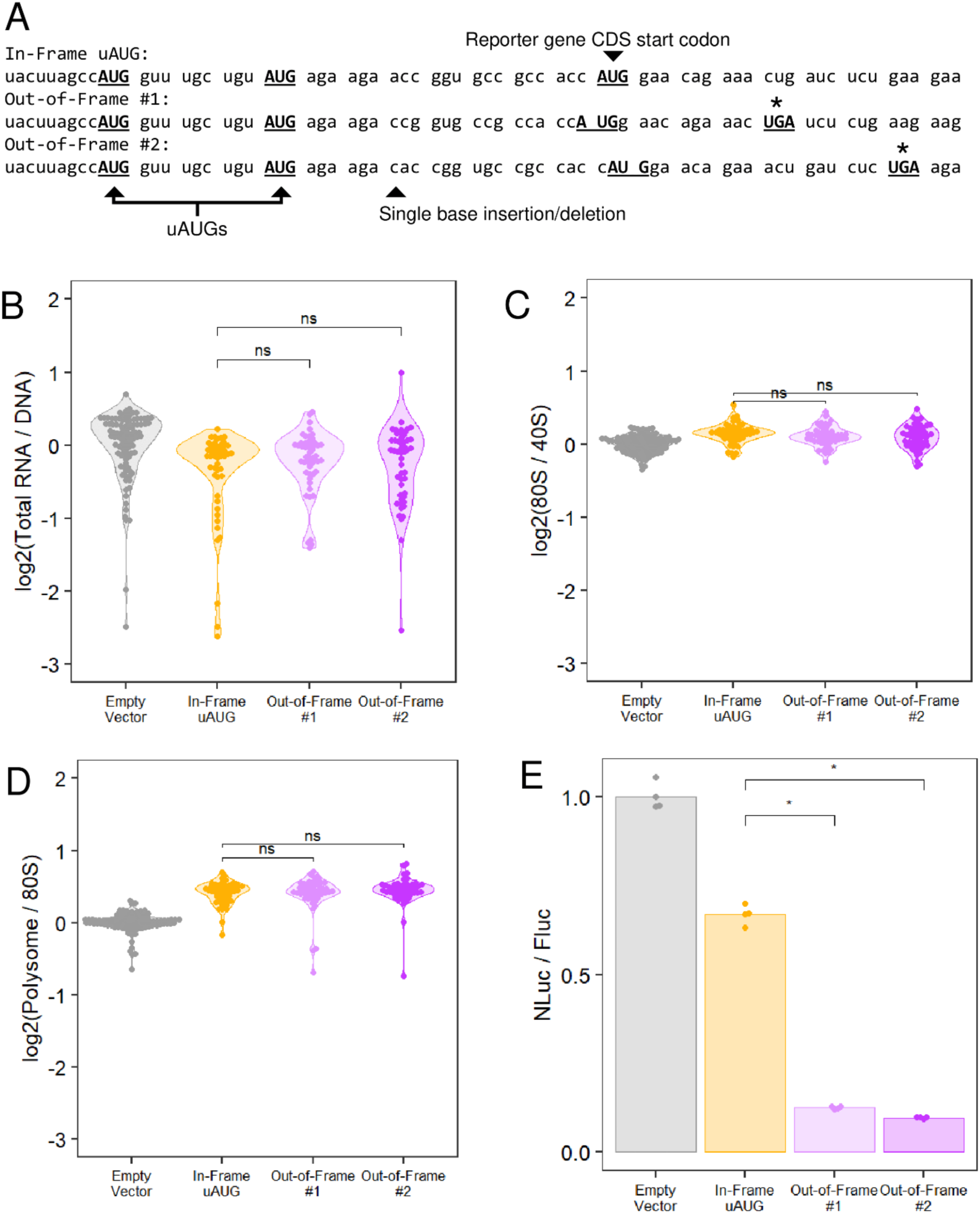
Out-of-frame ORF control sequences result in large decreases in reporter protein expression without showing an effect on transcript abundance or translational measures in the MPRA. **A)** A control sequence was designed with uAUG start codons in-frame with the reporter coding sequence, creating an oORF and N-terminal extension of the reporter gene. Two out-of-frame variants were generated by a single base insertion or deletion. **B-D)** Relative to the in-fame uAUG sequence, the two frame-shifted oORFs did not result in significant changes in transcript abundance, 80S/40S enrichment, or polysome/80S enrichment. **E)** These controls did however decrease luciferase reporter expression significantly (n=4). (Mann-Whitney U test: * p < 0.05)

**Figure S7:**
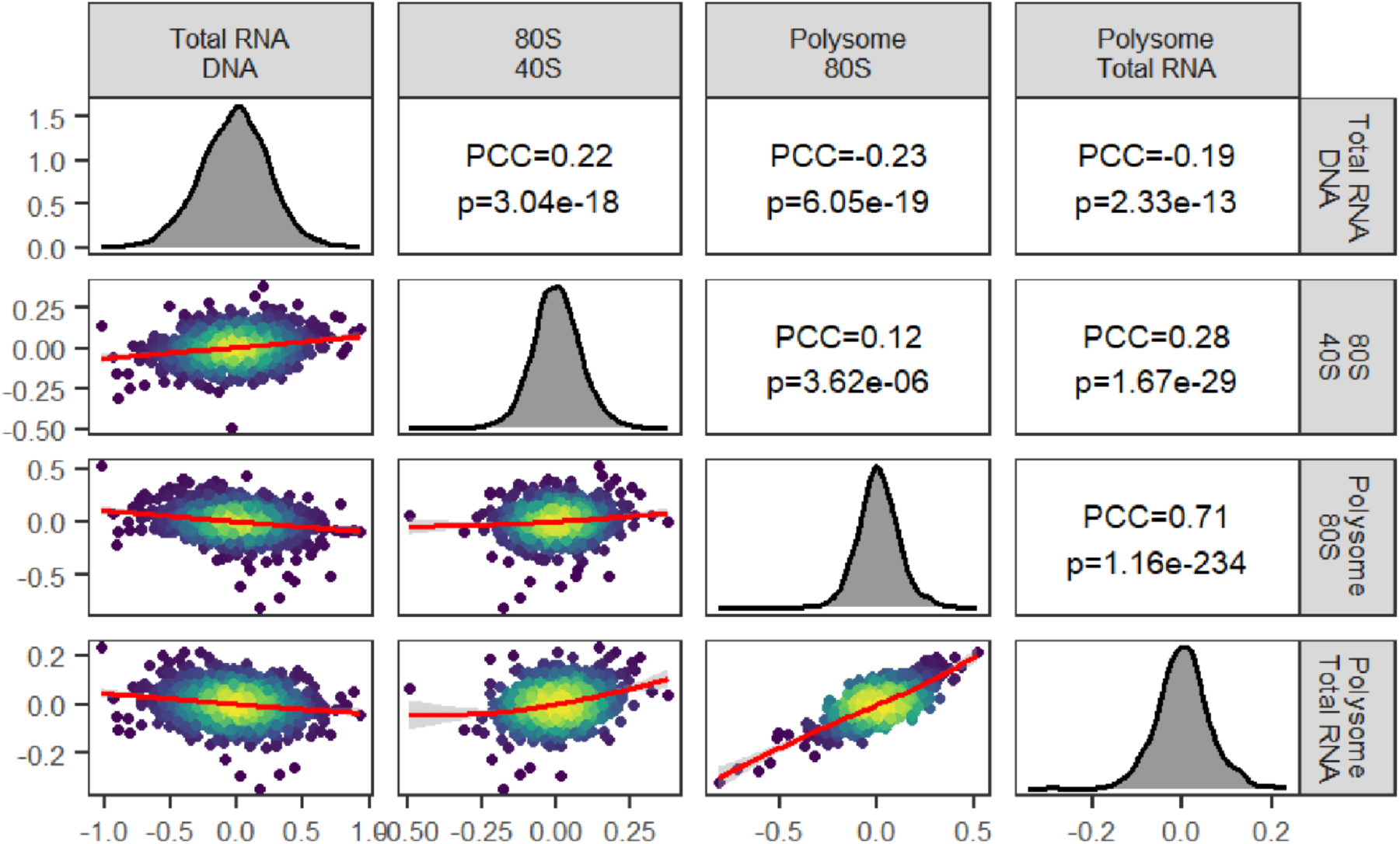
Allele log2-fold changes in transcript abundance and translational measures in HEK cells show little correlation. While polysome/80S and polysome/total RNA log2-fold changes were relatively well correlated, changes in 80S/40S enrichment were distinct.

**Figure S8:**
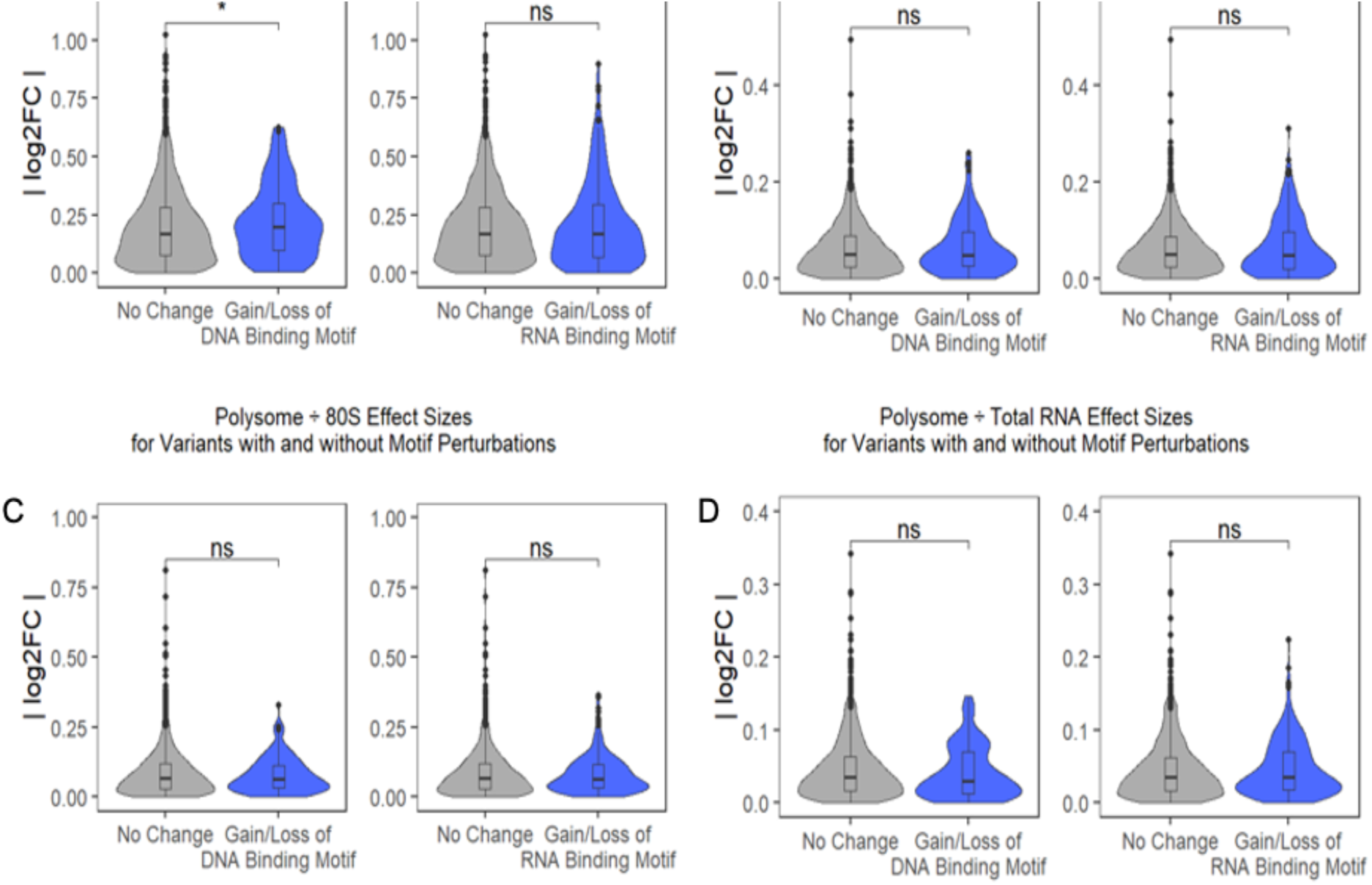
Variants that perturb predicted DNA or RNA binding motifs generally do not have larger effect sizes. **A)** Variants that result in the gain or loss of at least one DNA binding motif (Weirauch et al., 2014) have nominally greater effects on transcript abundance. **B-D)** However, changes in translational measures are not greater on average for variants that alter the presence of RNA-binding protein binding motifs (Ray et al., 2013). (Mann-Whitney U test: * p < 0.05)

**Figure S9:**
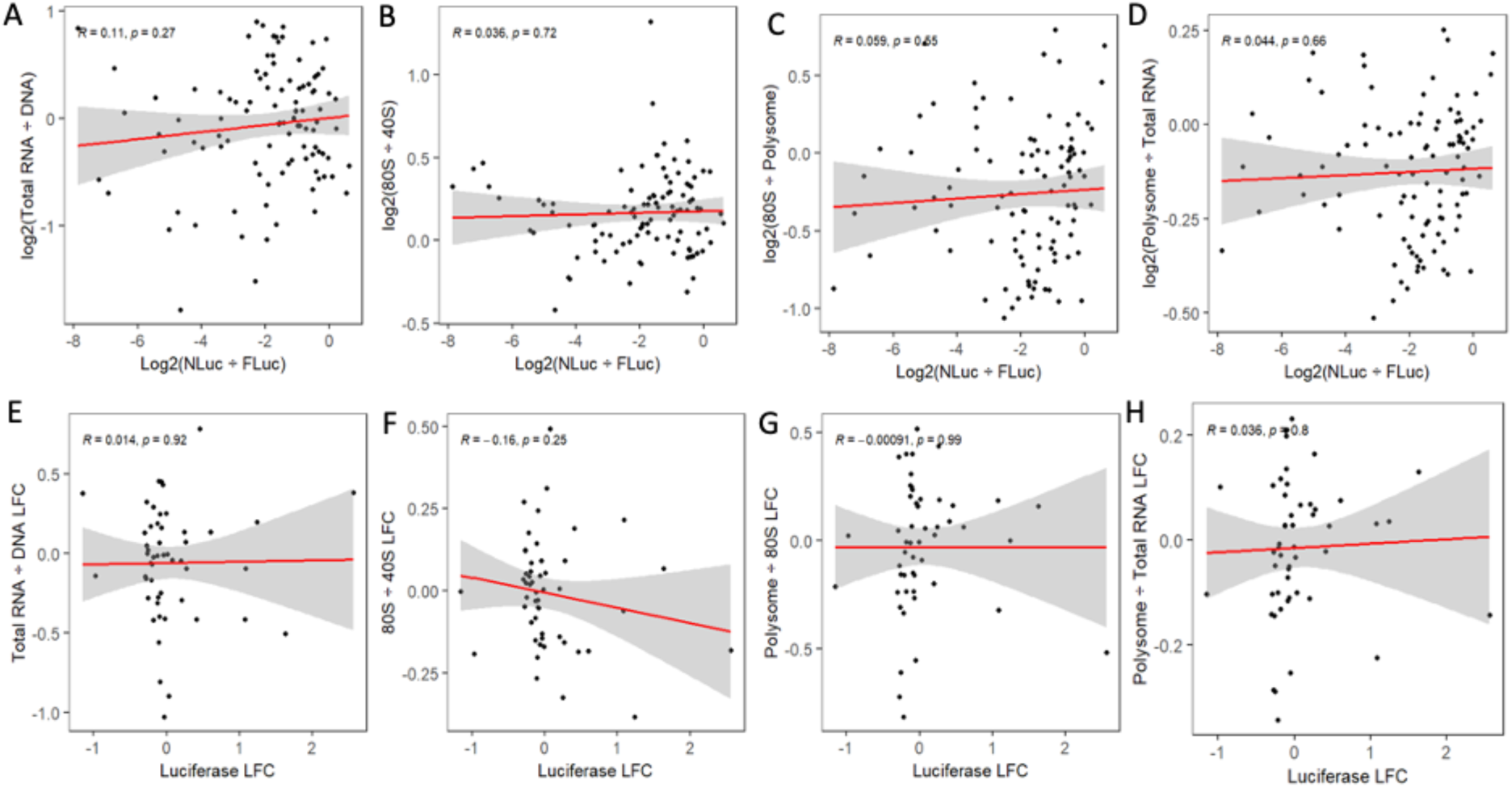
No single MPRA measurement predicts luciferase reporter gene expression or allelic effects on expression. **A-D)** The log2-luminescence ratio of 102 reporter constructs does not correlate directly with transcript abundance, 80S/40S enrichment, polysome/80S enrichment, or polysome/total RNA enrichment. **E-H)** Additionally, no correlation is found in the allelic log2-fold change for any of these measures.

**Figure S10:**
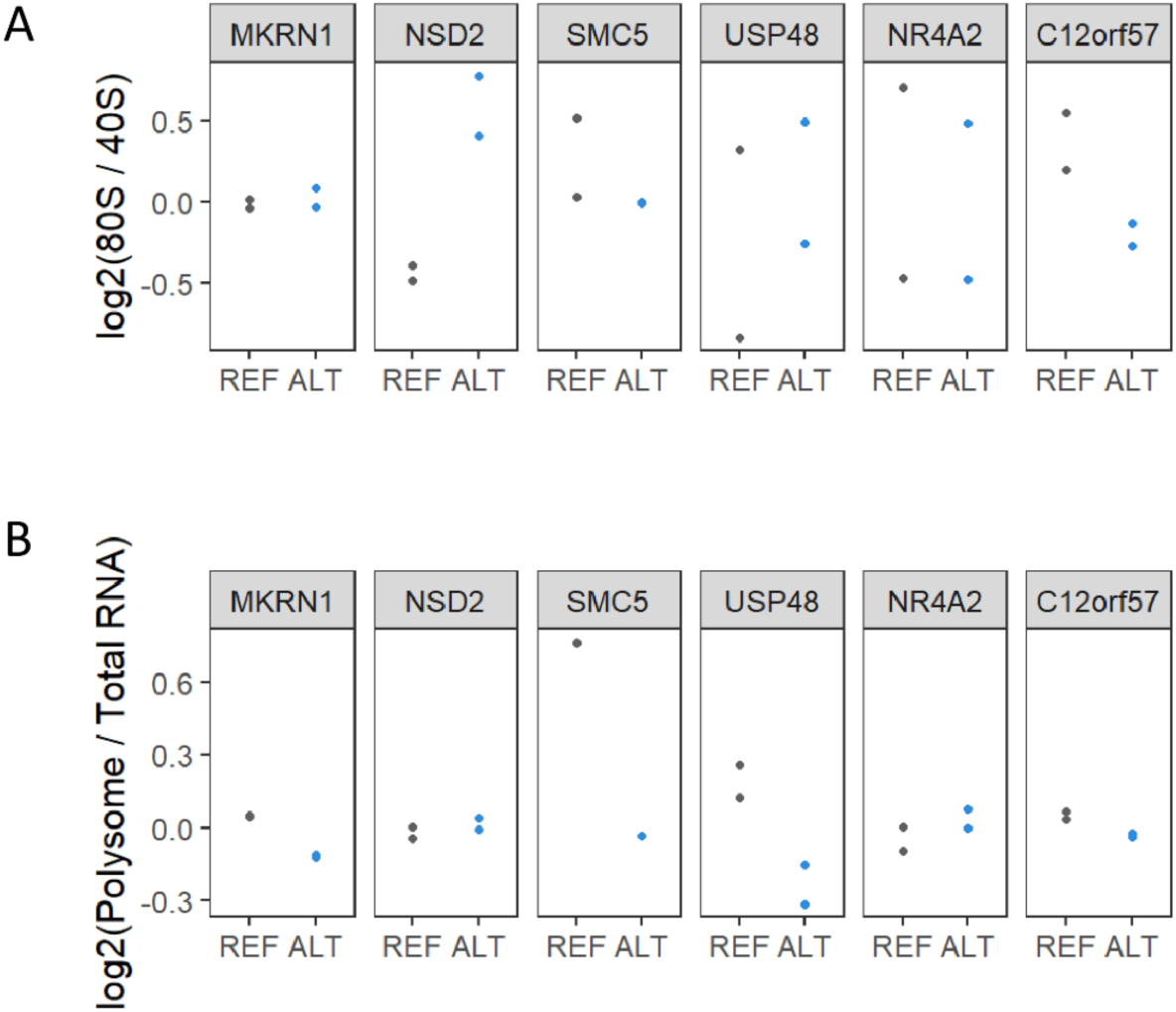
Proband variants alter relative polysome and monosome occupancy in the context of the native transcripts. **A)** Measurement of the relative allele abundance in 80S and 40S RNA fractions from proband-derived LCLs reveals an effect of proband variants in *NSD2* and *C12orf57*. **B)** Highly robust allelic effects in polysome/total RNA enrichment are found for proband variants in *SMC5* and *MKRN1*.

**Figure S11:**
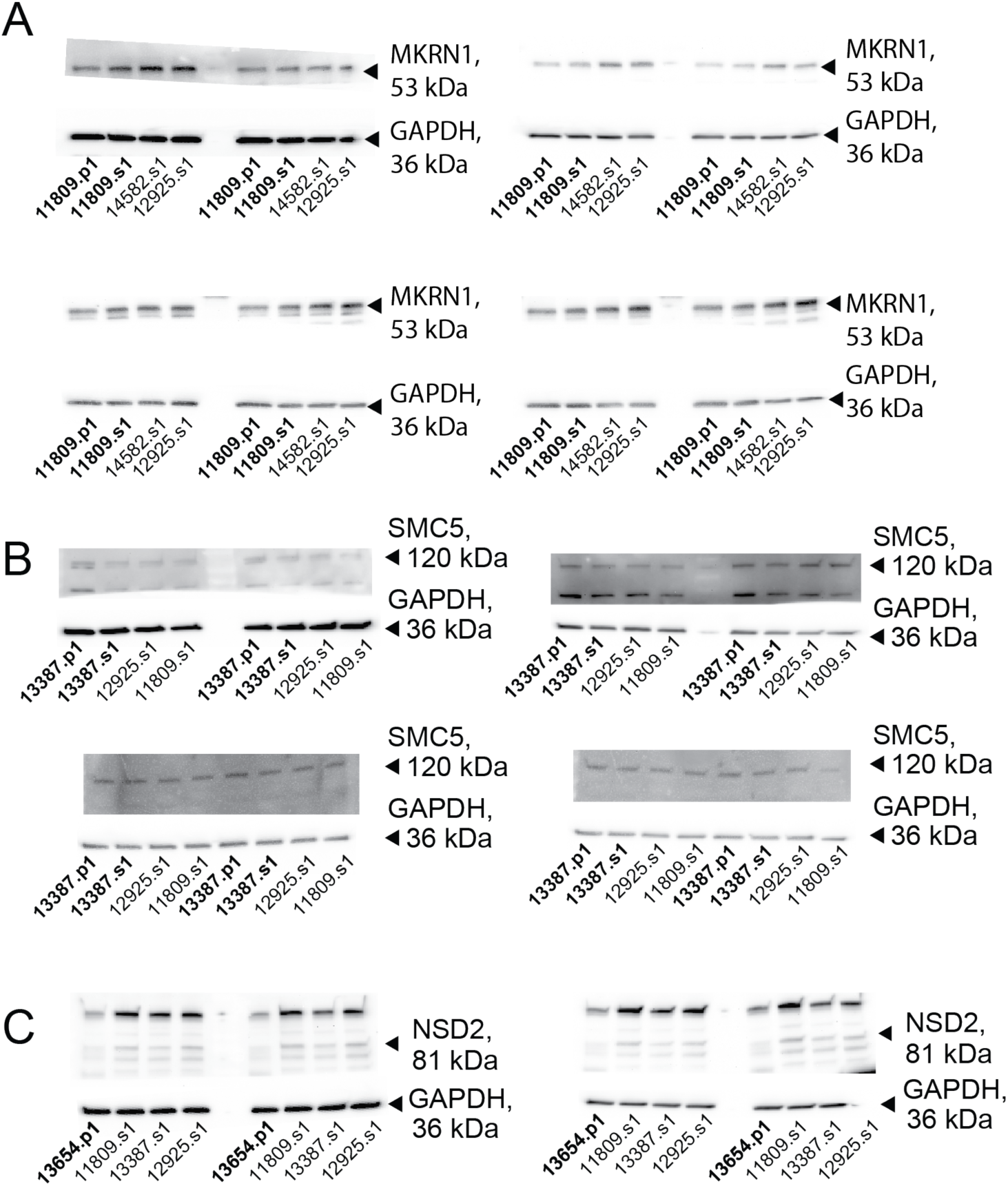
Some proband variants alter protein expression of the associated gene relative to matched sibling samples and unrelated samples. **A)** Western blots for MKRN1 were performed in 8 technical replicates with GAPDH as a loading control. Lanes marked 11809.p1 are LCL samples from the proband carrying the mutation in *MKRN1*, and lanes marked 11809.s1 are the matched sibling control LCL sample. Two additional unrelated non-proband samples were run as well. The bands used for quantification are denoted by the arrows. **B)** Western blots for SMC5 were performed in 8 technical replicates with GAPDH as a loading control. Lanes marked 13387.p1 and 13387.s1 are the matched proband and sibling, respectively. **C)** Western blots for NSD2 with GAPDH as a loading control were performed in 4 technical replicates. Lanes marked 13654.p1 are LCL samples from the proband carrying a mutation in *NSD2*. A matched sibling cell line was not available, and three unrelated non-proband controls were used.

**Figure S12:**
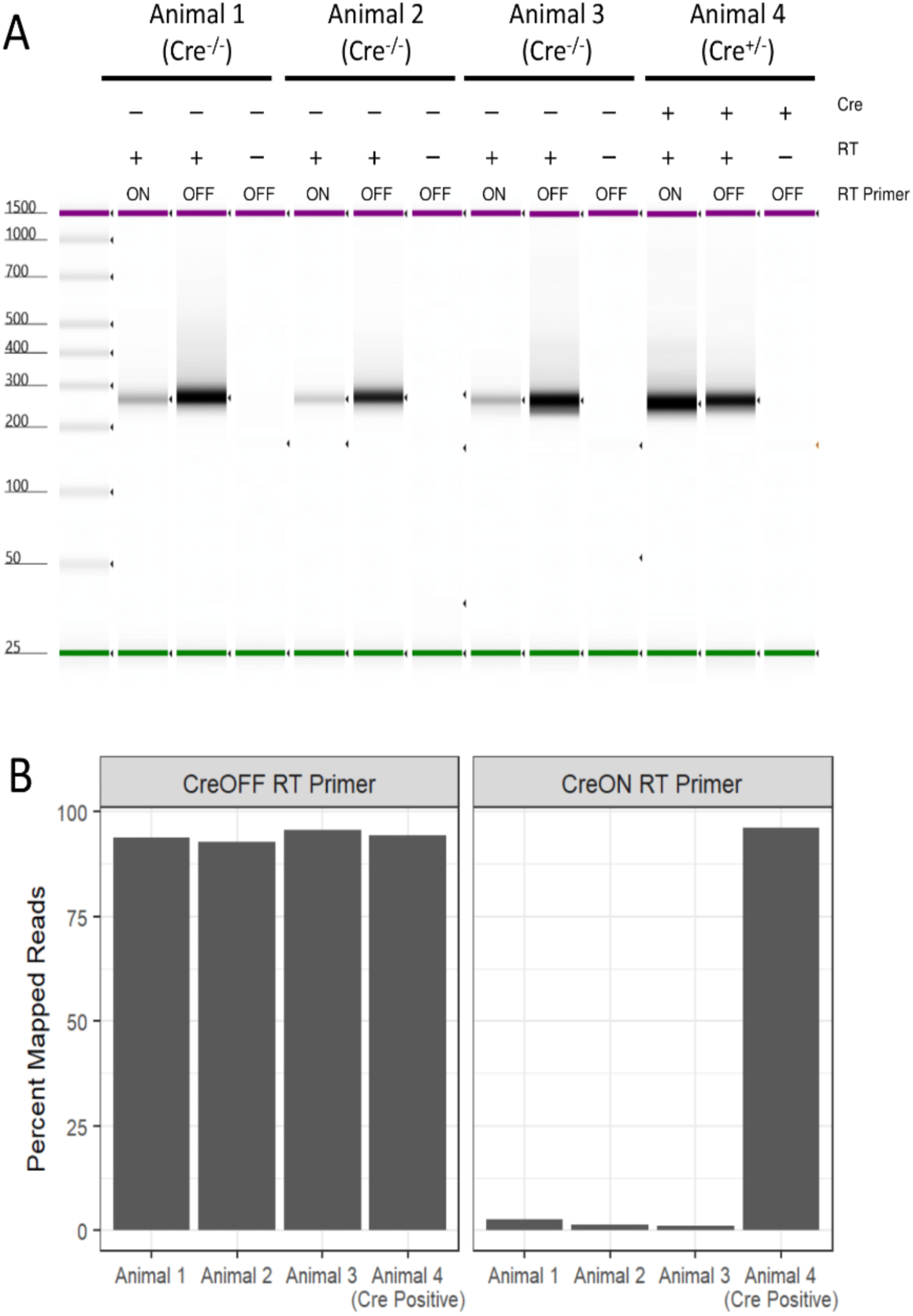
A Cre-invertible reverse transcription primer allows for selective amplification of mRNA from Cre-positive cells. **A)** Cre^+/-^ and Cre^-/-^ offspring from a cross between Ef1a:LSL-eGFP-RPl10 and Vglut1-IRES2-Cre-D were transduced with subpools of the MPRA library (sublibrary 1 in animal 1, sublibrary 2 in animal 2, sublibrary 3 in animal 3, and sublibrary 1 in animal 4). Total RNA extracted from the cortex of each animal at day P21 was subjected to the standard library preparation protocol but varying the reverse transcription (RT) primer used or the addition of Superscript IV reverse transcriptase. After indexing PCR, HSD1000 Tapestation ScreenTapes were used to measure library fragment size distribution and concentration. For all four animals, no final library product was measured without the addition of reverse transcriptase to the cDNA synthesis step. Libraries were generated for all four animals from cDNA primed with the CreOFF_RT_UMI (OFF) primer. Libraries were also generated for all four animals when using the CreON_RT_UMI (ON) primer; however, less product was generated in the final indexing PCR for the Cre^-/-^ animal libraries. **B)** Sequencing reads from libraries primed with CreOFF_RT_UMI mapped to the reporter construct and barcode sequences at similar rates (93-96%). The CreON_RT_UMI-primed libraries from Cre^-/-^ animal mapped at rates less than 3% (compared to 96% for animal 4) indicating that products amplified from Cre-negative animals were non-specific.

**Figure S13:**
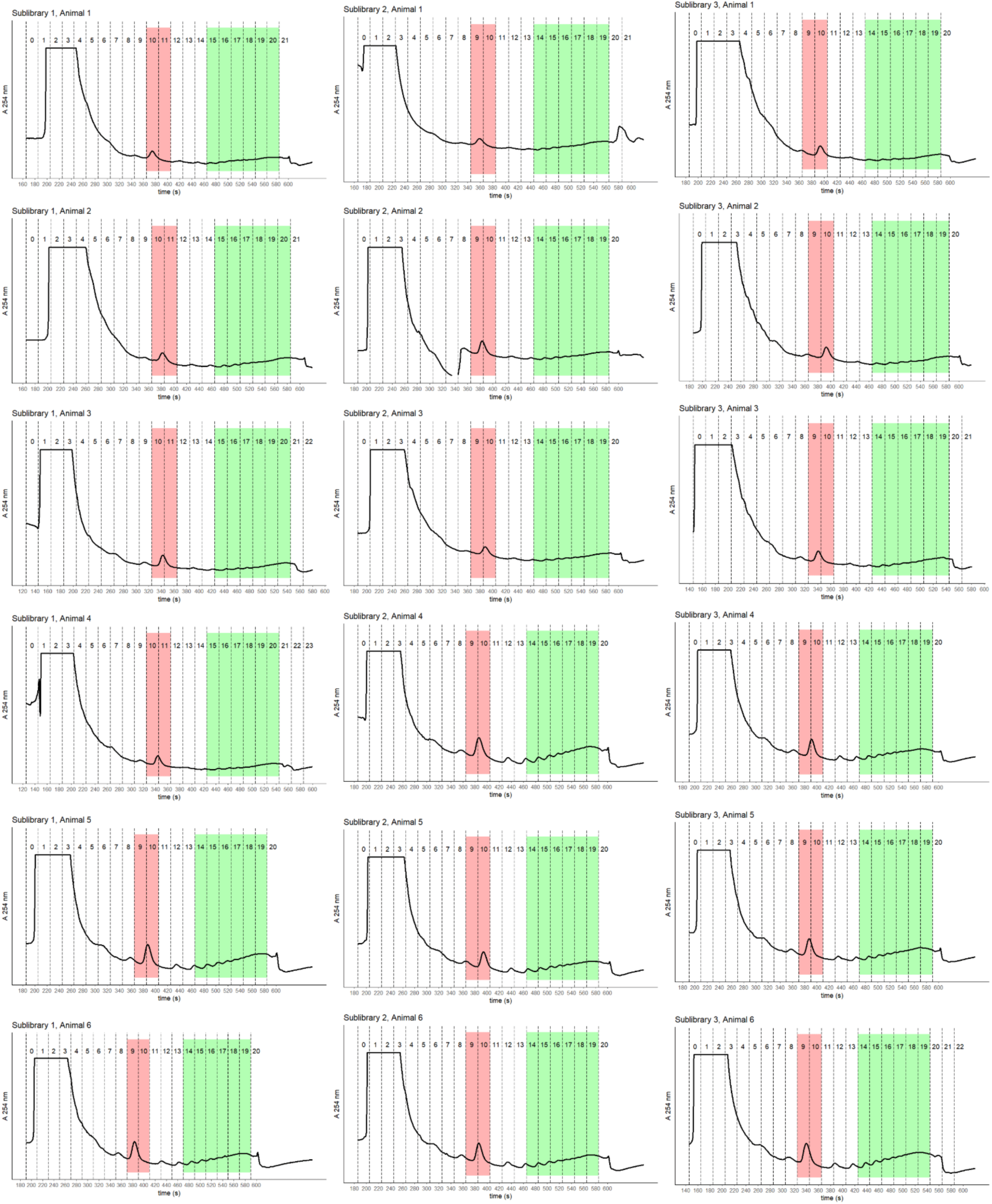
Polysome fraction traces for Vglut MPRA lysates. Lysates were prepared from homogenized cortical tissue taken from animals transduced with one of three MPRA sublibraries. Six replicate animals were used for each sublibrary. Lysates were fractionated after density centrifugation through 10-50% sucrose gradients by upward displacement, and absorbance at 254 nm was monitored. Monosome- and polysome-associated RNA were extracted from the fractions shaded.

**Figure S14:**
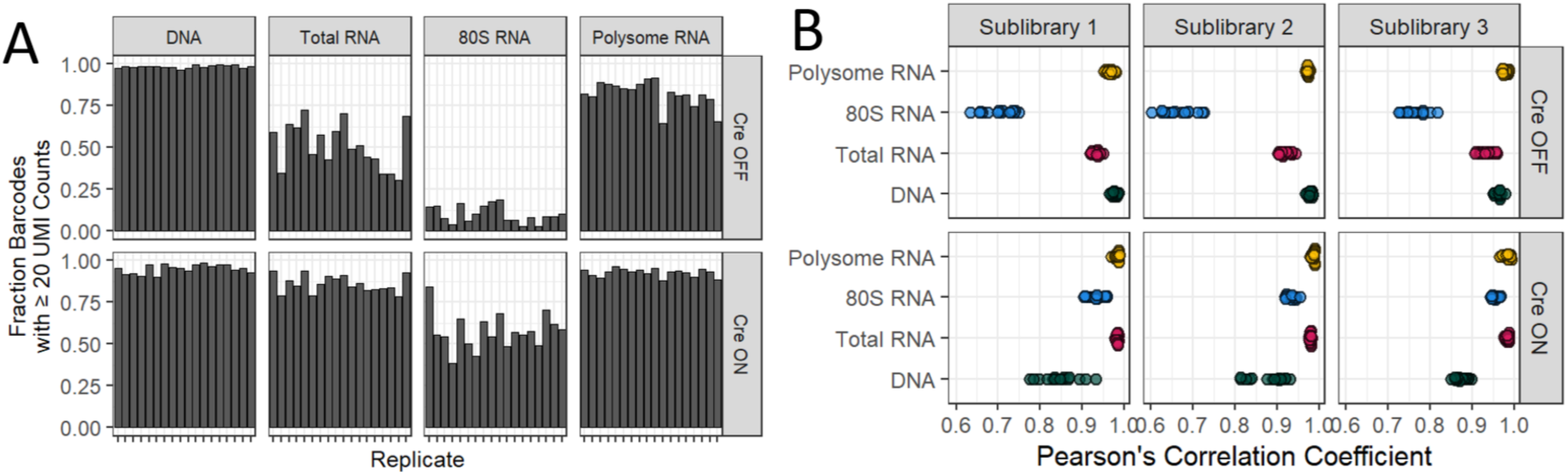
Barcode recovery rate and pairwise replicate correlations from Vglut MPRA libraries. **A)** The rate of barcode recovery with 20 or more UMI counts varied by the sequenced fraction and the Cre-state of the library. The 80S RNA libraries prepared with the Cre OFF RT primer had particularly low barcode recovery rates. **B)** All pairwise comparisons within each fraction type show a generally high correlation (Pearson’s correlation coefficient > 0.9) between replicates with Cre OFF 80S RNA and Cre ON DNA libraries being the exceptions.

**Figure S15:**
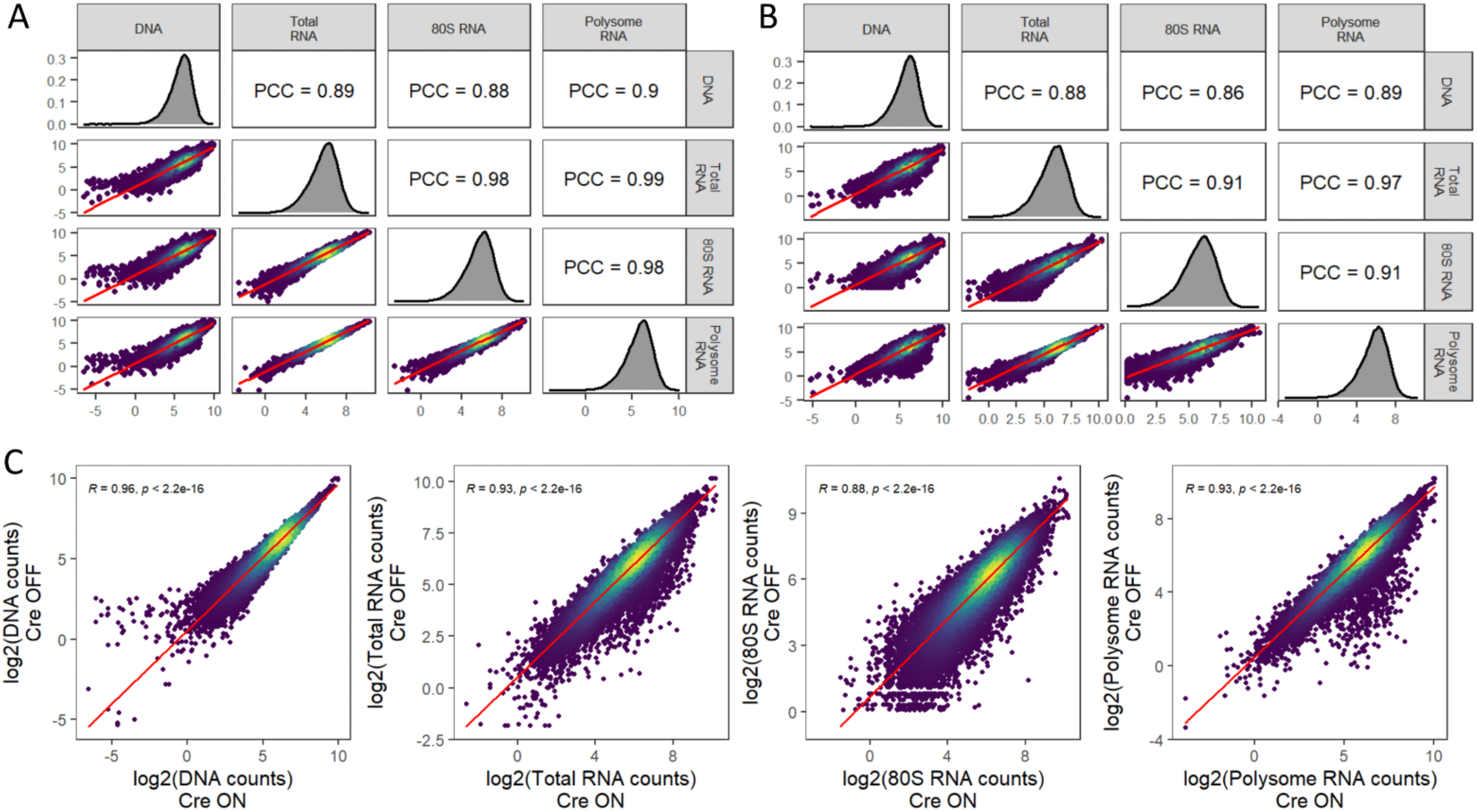
Correlations between *in vivo* measured DNA and RNA libraries and between Cre ON and Cre OFF. **A-B)** Barcode counts (averaged across animals and log2-transformed) show high correlation across both RNA and DNA libraries, with RNA libraries correlating more tightly together compared to DNA. **C)** The correlation of barcode counts (averaged across animals and log2-transformed) between Cre ON and Cre OFF is highest for DNA abundance with some divergence in barcode abundance across RNA fractions between the two cellular contexts.

**Figure S16:**
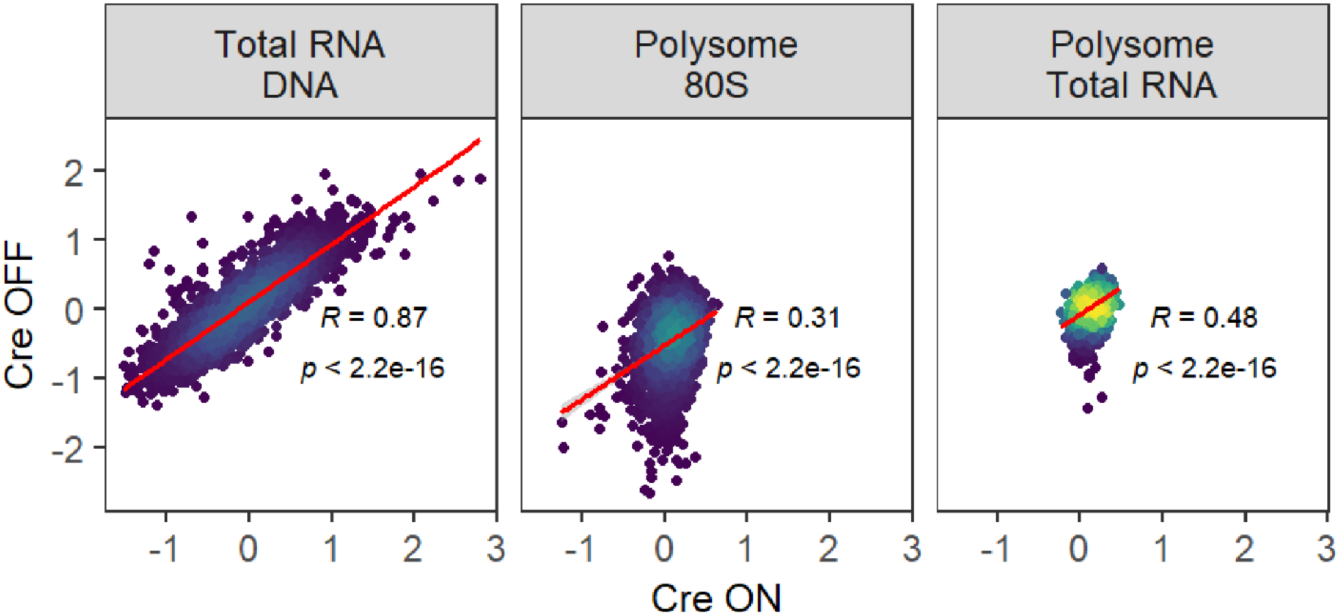
Correlations of *in vivo* transcript abundance and translational measures between Cre ON and Cre OFF cell types. Reporter transcript abundance [log2(total RNA / DNA)] measurements averaged across barcodes and animals show a high correlation across Cre ON and Cre OFF libraries. Measures of polysome/80S or polysome/total RNA enrichment span narrower ranges than transcript abundance in both cellular contexts and show significantly lower correlations.

**Figure S17:**
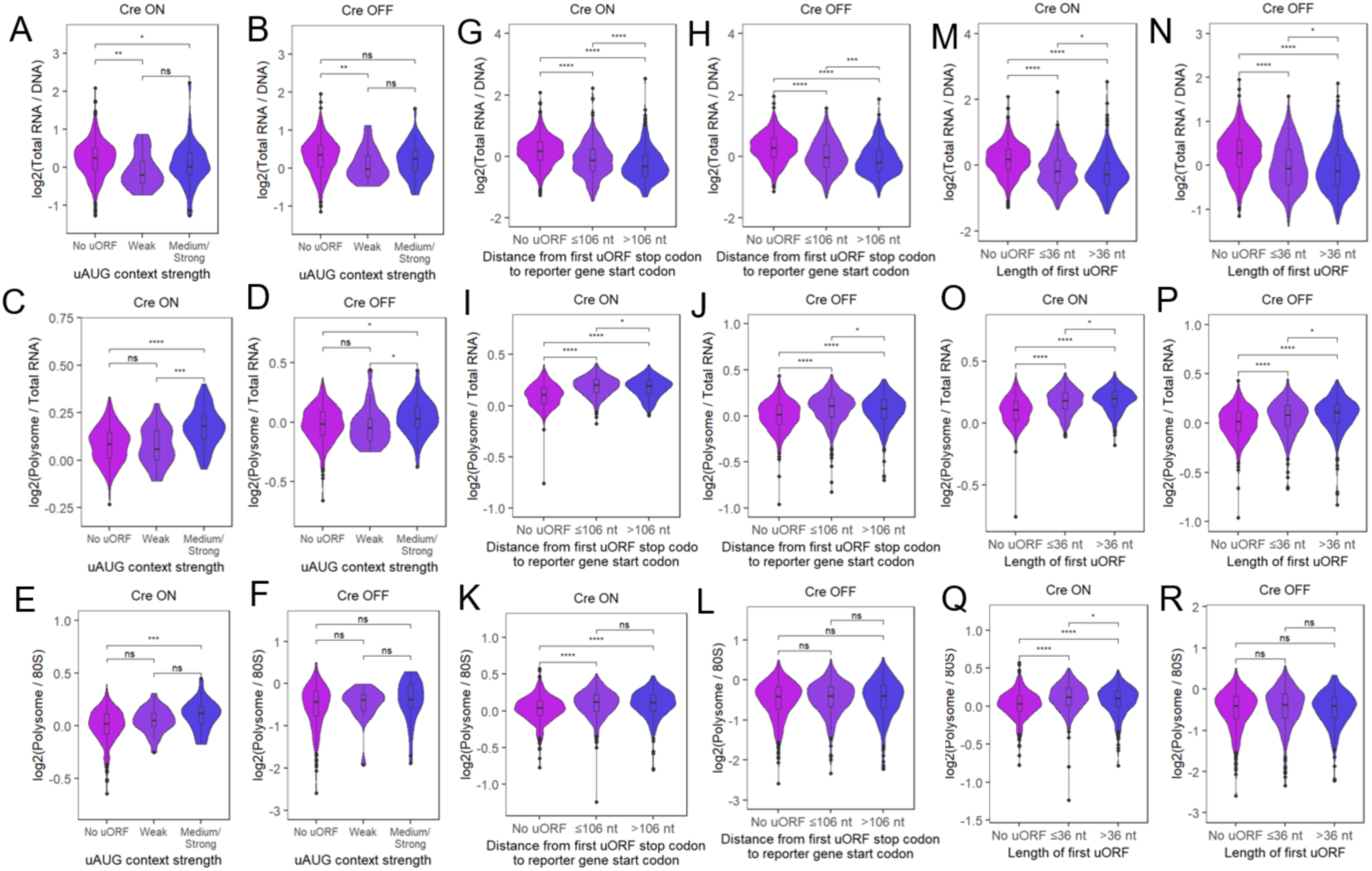
The *in vivo* effects of uORFs on transcript abundance and translation depend on the context of uORF start codons and the position of uORF stop codons within 5′UTR. **A-F**) In glutamatergic neurons (Cre ON), the effect uORFs have on translational measures (polysome/total RNA and polysome/80S enrichment) is greater when the uAUG context is more favorable. Less of an effect of uAUG context is observed for transcript abundance measurements. **G-L)** A greater distance between the stop codon of the first uORF in the UTR and the reporter gene (>106 nt) results in a stronger effect of the uORF on decreasing transcript abundance *in vivo*. This distance has a relatively smaller effect on translational measures. **M-R)** The length of the uORF has a small nominal effect on both transcript abundance and polysome enrichment *in vivo*. (Mann-Whitney U test: * p<0.05, ** p < 0.01, **** p < 0.0001)

**Figure S18:**
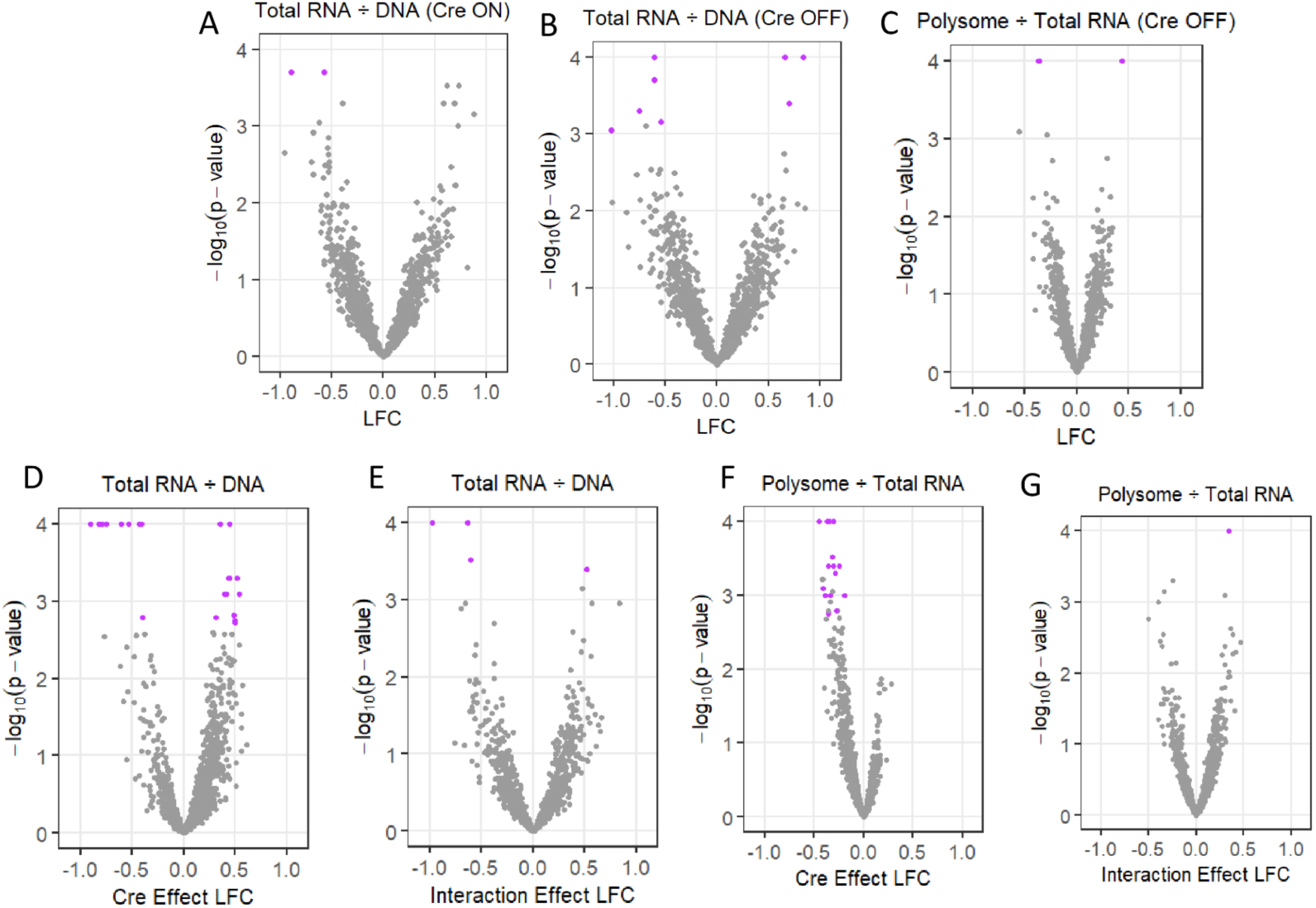
Variant effects discovered across Cre ON or Cre OFF conditions, and allelic effects are generally not affected by cellular context. **A-B)** Several variants had functional effects on transcript abundance in either glutamatergic neurons (Cre ON) or other brain cell types (Cre OFF), although only one variant was common across both conditions. **C)** Similarly, two variants impacting polysome/total RNA in Cre OFF cells were not discovered in glutamatergic neurons. **D-F)** By modeling the interaction of allelic effects with Cre-state, over a dozen reference alleles show some difference between Cre ON and Cre OFF. **E-G)** Variants that show a significant difference in allelic effect across Cre-state. (purple points indicate comparisons with q-value < 0.05)

**Figure S19:**
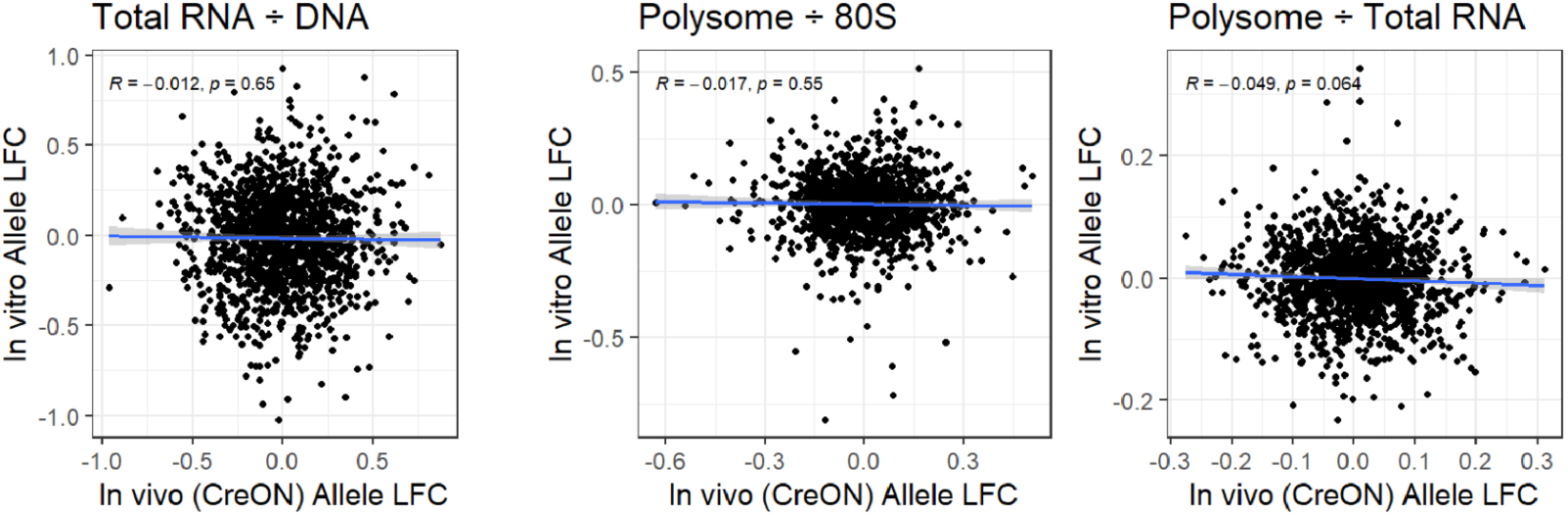
Variant effects do not correlate between the HEK MPRA and the *in vivo* MPRA measured in glutamatergic neurons. Correlation of HEK allelic effects (Y-axis) to *in vivo* allelic effects show little correlation.

